# A systematic scoping review of studies describing human factors, human-centered design, and usability of sensor-based digital health technologies

**DOI:** 10.1101/2024.02.23.24303220

**Authors:** Animesh Tandon, Bryan R Cobb, Jacob Centra, Elena Izmailova, Nikolay V. Manyakov, Samantha McClenahan, Smit Patel, Emre Sezgin, Srinivasan Vairavan, Bernard Vrijens, Jessie P Bakker, the Digital Health Measurement Collaborative Community hosted by the Digital Medicine Society (DiMe)

## Abstract

**Background:** Increasing adoption of sensor-based digital health technologies (sDHTs) in recent years has cast light on the many challenges in implementing these tools into clinical trials and patient care at scale across diverse patient populations; however, the methodological approaches taken towards sDHT usability evaluation have varied markedly.

**Objective:** To elucidate the current landscape of studies reporting data related to sDHT human factors, human-centered design, and/or usability.

**Methods:** We conducted a systematic scoping review of studies published between 2013 and 2023 and indexed in PubMed, in which data related to sDHT human factors, human-centered design, and/or usability were reported. Following a systematic screening process, we extracted the study design; participant sample; the sDHT(s) used; the method(s) of data capture; and the type(s) of usability-related data captured.

**Results:** Our literature search returned 442 papers, of which 85 were found to be eligible and 83 were available for data extraction and not under embargo. In total, 164 sDHTs were evaluated; 141 were wearable tools while the remaining 23 were ambient tools. The majority of studies (n=55; 66%) reported summative evaluations of final-design sDHTs. Almost all studies (n=82; 98%) captured data from targeted end-users, but only 18 (22%) captured data from additional users such as carepartners or clinicians. User satisfaction and ease of use were evaluated for >80% of sDHTs; however, learnability, efficiency, and memorability were reported for only 11 (13%), 4 (5%), and 2 sDHTs (2%), respectively. Fourteen sDHTs (17%) were evaluated according to the extent to which users were able to understand the clinical data or other information presented to them (understandability) and/or the actions or tasks they should complete in response (actionability). Notable gaps in reporting included the absence of a sample size rationale (reported for 25% of all studies and 31% of summative studies) and incomplete sociodemographic descriptive data (complete age, sex/gender, and race/ethnicity reported for 17% of studies).

**Conclusions:** Based on our findings, we suggest four actionable recommendations for future studies that will help to advance the implementation of sDHTs: 1) Consider in-depth assessment of technology usability beyond user satisfaction and ease of use; 2) Expand recruitment to include important user groups such as clinicians and carepartners; 3) Report the rationale for key study design considerations including the sample size; and 4) Provide rich descriptive statistics regarding the study sample to allow a complete understanding of generalizability to other patient populations and contexts of use.

## Introduction

Sensor-based digital health technologies (sDHTs), defined as connected digital medicine products that process data captured by mobile sensors using algorithms to generate measures of behavioral and/or physiological function [1], have been increasingly adopted in both research and healthcare in recent years [2,3]. Although regulatory guidance and a published framework focused on verification, analytical validation, and clinical validation processes for sDHTs have been widely adopted [1,4–6], detailed best practices focused on human factors, human-centered design, and/or usability (defined in Box 1) of sDHTs have not been clearly described. Given that sDHTs (A) encompass a wide spectrum of tools which may or may not meet the United States Food and Drug Administration (FDA) definition of a medical device [4]; (B) take various forms such as wearable, ingestible, implantable, and ambient tools [2]; and (C) are applicable to both clinical research and clinical practice [7], the methodological approaches taken towards sDHT usability evaluation have varied markedly [8,9].

Increasing adoption of sDHTs in recent years has cast light on the many challenges in implementing these tools into clinical trials and patient care at scale across diverse patient populations [10,11]. Optimal sDHT implementation requires integration into existing research and clinical workflows to be impactful, but a “one-size-fits-all” approach fails to address the needs of sDHT users - including but not limited to patients/participants, their carepartners, clinicians, and investigators - or the complexities of each healthcare system [12–15]. For example, the physical size of an sDHT may limit its deployment in children; those with limited dexterity may not be able to manipulate a wearable appropriately; and those with poor vision may be limited in their ability to read information presented on a screen[16,17], highlighting the importance of human-centered design which prioritizes the needs, capabilities, and behaviors of users during the design process [18]. Inadequate attention to human-centered design and usability testing approaches can hinder the evaluation of healthcare interventions, contribute to insufficient adoption, perpetuate health disparities, increase costs, and potentially introduce safety risks [19–22]. Thus, integrating human factors considerations in the design, development, and evaluation of sDHTs is critical to improve their likelihood of being adopted and properly utilized in clinical research and healthcare in a way that is safe, effective, inclusive, and optimizes the user experience.

Recognizing the urgency of addressing these challenges, a pre-competitive collaboration within the Digital Health Measurement Collaborative Community (<u>DATAcc</u>) hosted by the Digital Medicine Society (<u>DiMe</u>) undertook a systematic scoping review examining the methodological approaches employed in published studies undertaken in human participants which reported data related to sDHT human factors, human-centered design, and/or usability. Our objective was to elucidate the current landscape and identify gaps, which will inform development and dissemination of recommendations and an evaluation framework of sDHTs as being fit-for-purpose from a usability perspective.

### Box 1

**Definitions**

#### Human factors

The application of knowledge about human behavior, abilities, limitations, and other characteristics of users to the design and development of an sDHT to optimize usability within a defined intended use or context of use. This definition incorporates terminology and concepts from the United States Food and Drug Administration [23], the United Kingdom Medicines and Healthcare products Regulatory Agency (MHRA) [24], and the National Medical Products Administration (NMPA) of China [translated] [25].

#### Human-centered design

An approach to interactive systems that aims to make systems usable and useful by focusing on the users, their needs and requirements, and by applying human factors and usability knowledge and techniques, as defined in the International Organization for Standardization (ISO) 9241-210:2019 standard [18].

#### Usability

The extent to which an sDHT can be used to achieve specified goals with ease, efficiency, and user satisfaction within a defined intended use or context of use. This definition incorporates terminology and concepts from the FDA [23], the MHRA [24], the NMPA [translated] [25], and ISO 9241-210:2019 [18].

## Methods

We followed the PRISMA guidelines for scoping reviews [26]. As a scoping review, this work did not meet the criteria for registration on PROSPERO [27]. The protocol is available from the corresponding author.

### Literature search

We completed our literature search in PubMed using search terms designed in six layers as follows (terms within each layer were separated by the Boolean operator “or”, while the layers themselves were separated using “and” or “not”): (A) Medical Subject Heading (MeSH; [28]) term for human participants; (B) MeSH terms related to sDHTs, such as *wearable electronic devices* and *digital technology*; (C) keywords related to sDHTs such as *wear\** [asterisk indicates truncation]*, remote,* and *connected*; (D) keywords related to human-centered design, usability, human factors, and ergonomics; (E) exclusion of out-of-scope publication types, such as editorials and case reports; and (F) published between January 1st 2013 and May 30th 2023. The complete search string is provided in Supplementary Table 1.

To avoid potentially overlooking novel or emerging technologies, the search terms did not include descriptions of specific sensor types (such as *accelerometer*), form factors (such as *watch*), methodology (such as *actigraphy*), wear location (such as *wrist*), or technology make/model.

### Study selection

We systematically screened publications identified in the literature search based on the PICO (patients/participants; intervention; comparator; outcomes) eligibility criteria outlined in Table 1, designed to identify studies describing the incorporation of knowledge about human behavior, abilities, limitations, and other characteristics of users to the design and development process; human-centered design; and/or ease of use, efficiency, and/or user satisfaction of sDHTs. Studies reporting sDHT usage/adherence (such as average wear-time) and/or measurement success metrics (such as percentage of in-range measurements obtained) were out of scope unless also reporting one of the aforementioned concepts.

**Table 1:** Study selection eligibility criteria.

| PICO Framework <sup>a</sup> | Eligibility criteria |
| --- | --- |
| Patient / participant | 1. Exclude studies that do not report data collected from human participants. |
| Intervention | 2. Exclude studies that do not assess a specific sDHT, defined according to the definition of BioMeT in the V3 Framework <sup>b</sup>:<br>a. Connected<br>Interpreted as a digital method of data transfer from the sDHT to the location of data analysis, either wired or wireless.<br>b. Mobile<br>Interpreted as the tool being capable of collecting data in the out-of-clinic setting, although the study may have deployed the tool in-clinic.<br>c. Sensor-based<br>Interpreted as the tool containing at least one sensor sampling a physical construct such as acceleration, light, or temperature.<br>d. Used for purposes of measurement, diagnosis, and/or treatment of a behavioral or physiological function. |
| Comparator | Not applicable. |
| Outcome/s | 3. Exclude studies that do not report data on human factors, human-centered design, or usability (see Box 1 for definitions). |

<sup>a</sup> The PICO framework is described in Eriksen et al. (2018, [31]).
<sup>b</sup> Throughout this review we refer to 'sensor-based digital health technology' (sDHT); however, this was operationalized according to the definition for 'biometric monitoring technology' (BioMeT) as described in Goldsack et al. (2020; [1]).

Two independent investigators (JC, JB) began by screening a random selection of 20% of publications; disagreements were resolved by consensus, and clarifications were made to the wording of the eligibility criteria to reduce ambiguity. The same two investigators then reviewed another random selection of 20% of publications; it was determined *a priori* that if the reviewers were in agreement for ≥90% of these publications, the remaining 60% would be reviewed by a single investigator (JC) as described elsewhere [29,30].

### Data extraction and analysis

Data extraction fields included study design and sample characteristics; the type, maturity, make/model, form factor, and wear location (if applicable) of each sDHT evaluated along with the health concept/s generated by each sDHT; the methodological approaches; and the types of usability-related data reported in each study. Most fields for data capture were categorical, with categories created in advance to minimize error.

Categories of usability-related data are described in Table 2, and compiled based on the literature including the International Standards Organization *Ergonomics of human-system interaction Part 210 Human-centered design for interactive systems* ISO 9241-210:2019 [18] and Nielsen’s (1994) usability attributes [32], as well as the studies identified in this review; that is, data not clearly fitting into an existing category were extracted and categorized post-hoc. We acknowledge that there are various models for capturing data describing usability and related topics [33]; however, there is no single standard that has been widely adopted.

**Table 2:** Categories of usability-related data extracted from eligible papers.

| Category | Definition |
| --- | --- |
| User satisfaction | The extent to which a user finds the sDHT to be pleasant to use, which may reflect trust, comfort, aesthetics, engagement, desirability, emotional response/s, and other considerations. Always captured through self-report. |
| Ease of use | The ease with which a user is able to perform user tasks. Can be captured through self-report (such as the mental demand or effort required to complete a task) or objective measures (such as the number of actions, number of attempts, or time required to complete a task). |
| Efficiency | The ease with which a user is able to perform user tasks after having learned how to use the sDHT. Captured according to the definition of ease of use above. |
| Learnability <sup>b</sup> | The ease with which a user is able to perform user tasks during their first encounter with the sDHT. Captured according to the definition of ease of use above. |
| Memorability | The ease with which a user is able to perform user tasks after a period of non-use, assessed in a test-retest paradigm. Captured according to the definition of ease of use above. |
| Usefulness <sup>c</sup> | The extent to which a user finds the sDHT, or its specific features/functions, to be valuable, productive, and/or helpful. Always captured through self-report. |
| Use-errors | An action or lack of action which may result in a use-related hazard (a potential source of harm), as well as error recovery defined as the ability of a user to make a correction following a use-error in order to complete a task. Can be captured through self-report or objective assessments. |
| Technical performance or malfunctions | Technical performance such as page load times, or the number, type, and severity of errors associated with sDHT malfunction. Can be captured through self-report or objective assessments. |
| Readability | The reading skills a user must possess to understand information presented to them through the sDHT itself, or through written materials such as instructions for use, cautions, warnings, and/or contraindications; [34]. Always captured through objective assessments, and typically reported as a reading grade. |
| Understandability and/or actionability | The extent to which users of diverse backgrounds, languages, and varying levels of health literacy understand (A) the clinical data or other information, such as instructions, cautions, warnings, and contraindications, presented to them; and (B) the actions or tasks they should complete in response, such as an sDHT-derived blood glucose measurement requiring an adjustment to medication [34]. Always captured through objective assessments. |

Study outcomes shown below the line are not typically considered usability data, but are related concepts often captured during usability evaluations.
Note that in the above definitions, 'self-report' includes data captured through surveys, interviews, and focus groups, while 'objective' includes data captured through observation (direct or video) or through the sDHT itself (or any related software), such as timestamps, app crash reports, and page load times.
<sup>a</sup> Comfort and trust were extracted separately for the purposes of this review.
<sup>b</sup> Learnability refers to operation of the sDHT rather than a practice effect associated with a research study outcome/endpoint.
<sup>c</sup> We have adopted the term usefulness instead of utility, to avoid confusion with clinical utility which refers to the extent to which implementing a medical product leads to improved health outcomes or provides useful information about diagnosis, treatment, management, or prevention of disease [35].

Consistent with the goal of a scoping review, all data were analyzed descriptively.

## Results

### Literature search and study selection

The PubMed search conducted on June 1st 2023 yielded 442 results, including one published only as an abstract. After applying the eligibility criteria described in Table 1, a further 356 publications were excluded. As such, 85 studies were determined to be eligible; however, two were under embargo, leaving 83 studies for data extraction (see Figure 1). A complete list of all included studies is provided in Supplementary Table 2.

**Figure 1:**
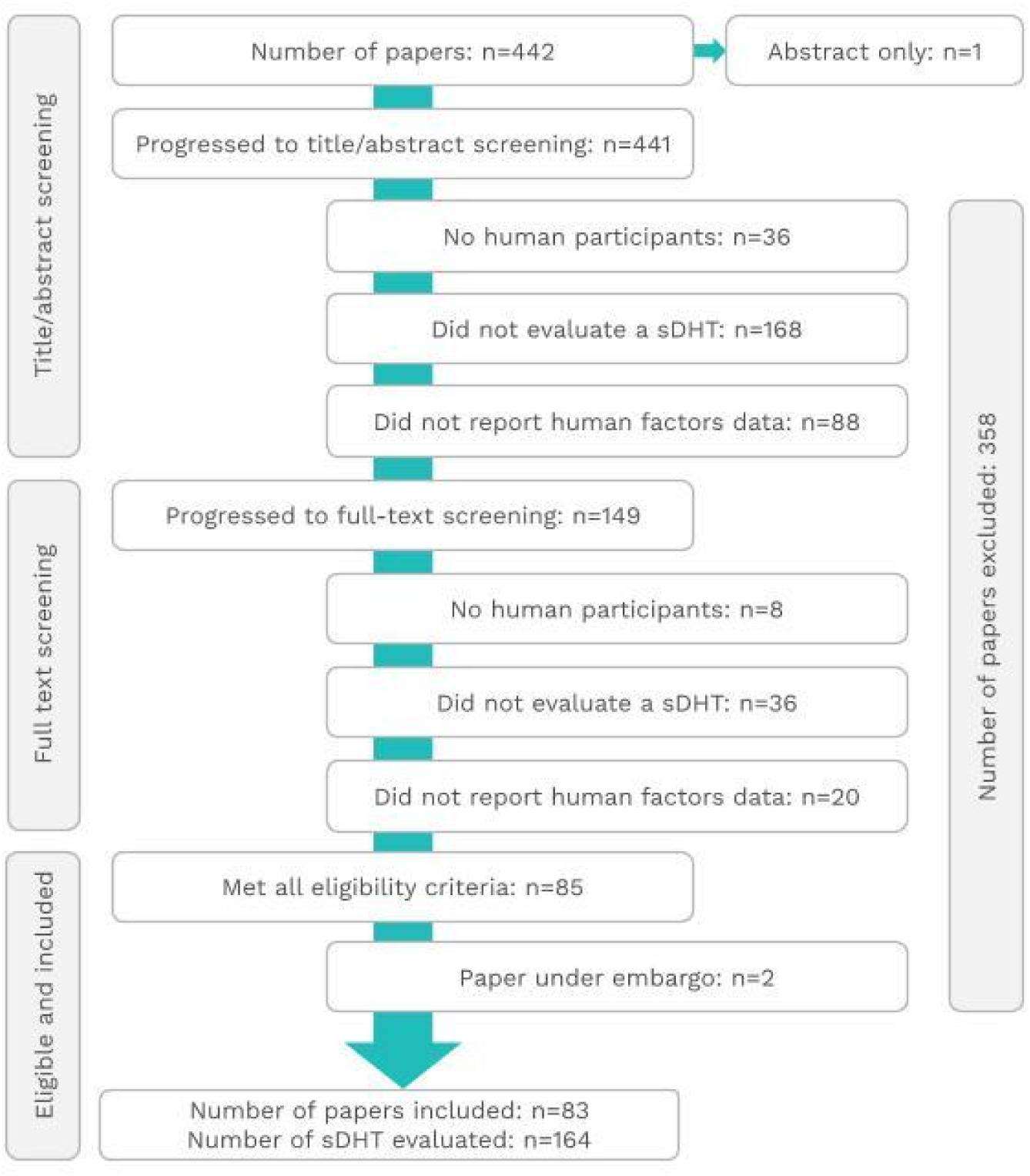
PRISMA (Preferred Reporting Items for Systematic Reviews and Meta-Analyses) flowchart

As described above, two investigators reached consensus on 20% of publications (88 of 442) before any further publications were screened. The same investigators then screened a further 88 publications independently, which resulted in 100% agreement of eligibility. Per protocol, a single investigator screened the remaining 266 papers.

### Study design considerations

The majority of studies (n=55; see Table 3) reported summative evaluations of products that were marketed or production-equivalent (that is, sample products of final design assembled in a way that differs from - but is equivalent to - the manufacturing processes used for the marketed product [36]). The remaining 28 studies reported formative evaluations of prototype products; we did not identify any reports focused solely on sDHT design. Most studies (n=53) were conducted partially or completely off-site. Study sample sizes spanned a wide range (*n*=1 to 623 with a median of n=27); however, only 21 of the full set of 83 studies (25%), and 17 of the 55 summative studies (31%), reported a rationale for the sample size (with or without a power calculation).

**Table 3:**
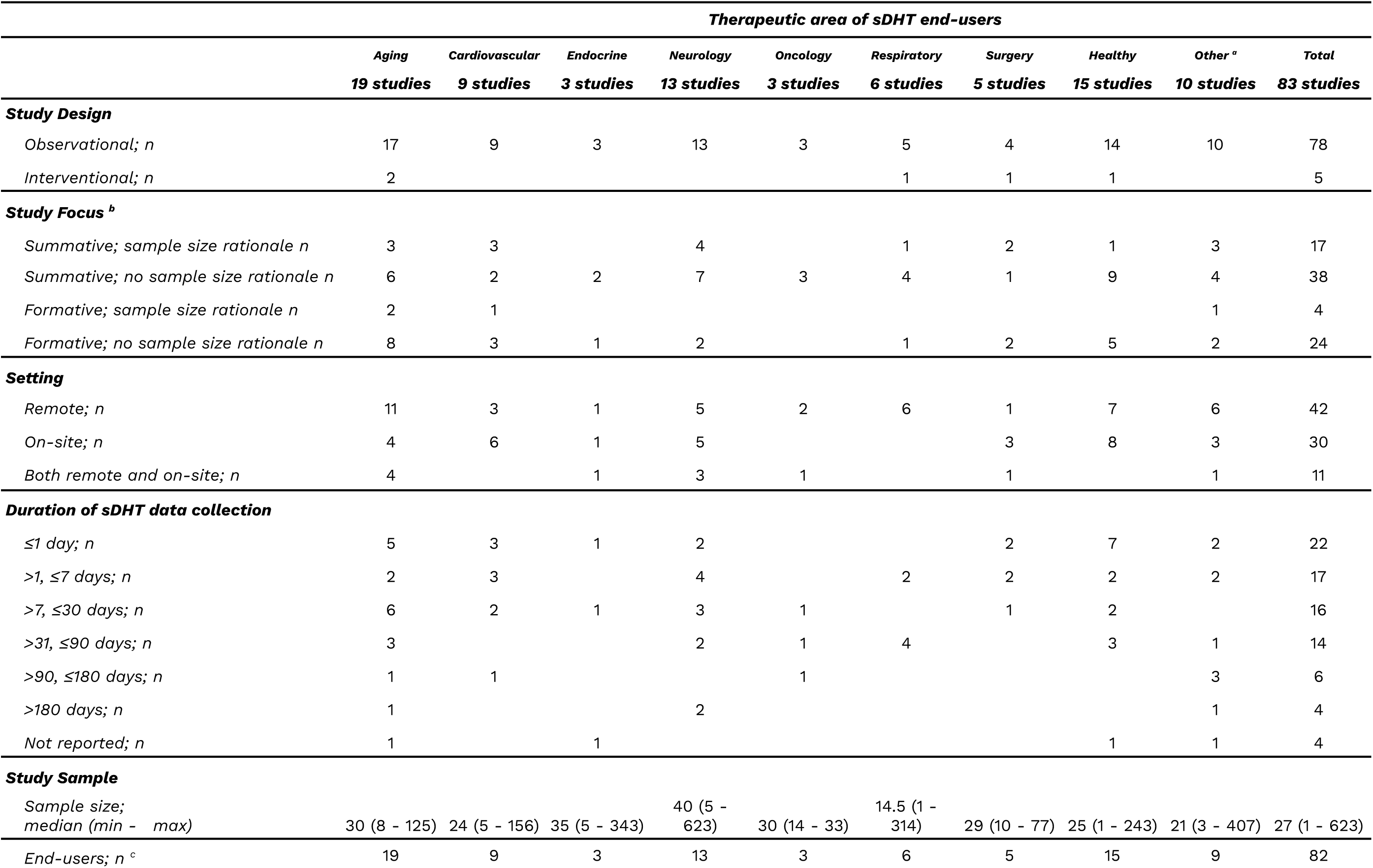
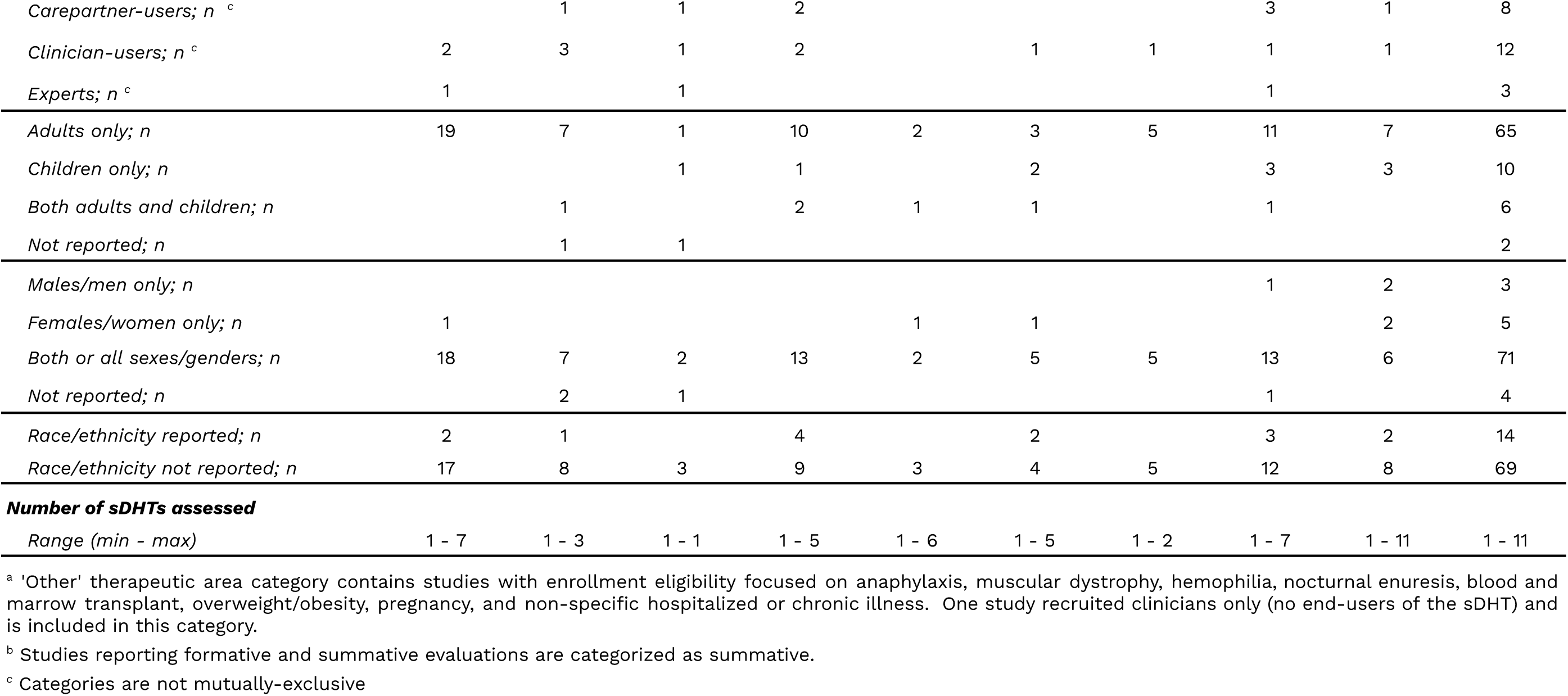
Study design and sample characteristics across therapeutic areas.

|  | Therapeutic area of sDHT end-users |  |  |  |  |  |  |  |  |  |
| --- | --- | --- | --- | --- | --- | --- | --- | --- | --- | --- |
|  | Aging<br>19 studies | Cardiovascular<br>9 studies | Endocrine<br>3 studies | Neurology<br>13 studies | Oncology<br>3 studies | Respiratory<br>6 studies | Surgery<br>5 studies | Healthy<br>15 studies | Other <sup>a</sup><br>10 studies | Total<br>83 studies |
| **Study Design** |  |  |  |  |  |  |  |  |  |  |
| Observational; n | 17 | 9 | 3 | 13 | 3 | 5 | 4 | 14 | 10 | 78 |
| Interventional; n | 2 |  |  |  |  | 1 | 1 | 1 |  | 5 |
| **Study Focus <sup>b</sup>** |  |  |  |  |  |  |  |  |  |  |
| Summative; sample size rationale n | 3 | 3 |  | 4 |  | 1 | 2 | 1 | 3 | 17 |
| Summative; no sample size rationale n | 6 | 2 | 2 | 7 | 3 | 4 | 1 | 9 | 4 | 38 |
| Formative; sample size rationale n | 2 | 1 |  |  |  |  |  |  | 1 | 4 |
| Formative; no sample size rationale n | 8 | 3 | 1 | 2 |  | 1 | 2 | 5 | 2 | 24 |
| **Setting** |  |  |  |  |  |  |  |  |  |  |
| Remote; n | 11 | 3 | 1 | 5 | 2 | 6 | 1 | 7 | 6 | 42 |
| On-site; n | 4 | 6 | 1 | 5 |  |  | 3 | 8 | 3 | 30 |
| Both remote and on-site; n | 4 |  | 1 | 3 | 1 |  | 1 |  | 1 | 11 |
| **Duration of sDHT data collection** |  |  |  |  |  |  |  |  |  |  |
| ≤1 day; n | 5 | 3 | 1 | 2 |  |  | 2 | 7 | 2 | 22 |
| >1, ≤7 days; n | 2 | 3 |  | 4 |  | 2 | 2 | 2 | 2 | 17 |
| >7, ≤30 days; n | 6 | 2 | 1 | 3 | 1 |  | 1 | 2 |  | 16 |
| >31, ≤90 days; n | 3 |  |  | 2 | 1 | 4 |  | 3 | 1 | 14 |
| >90, ≤180 days; n | 1 | 1 |  |  | 1 |  |  |  | 3 | 6 |
| >180 days; n | 1 |  |  | 2 |  |  |  |  | 1 | 4 |
| Not reported; n | 1 |  | 1 |  |  |  |  | 1 | 1 | 4 |
| **Study Sample** |  |  |  |  |  |  |  |  |  |  |
| Sample size;<br>median (min - max) | 30 (8 - 125) | 24 (5 - 156) | 35 (5 - 343) | 40 (5 - 623) | 30 (14 - 33) | 14.5 (1 - 314) | 29 (10 - 77) | 25 (1 - 243) | 21 (3 - 407) | 27 (1 - 623) |
| End-users; n <sup>c</sup> | 19 | 9 | 3 | 13 | 3 | 6 | 5 | 15 | 9 | 82 |
| *Carepartner-users; n<sup>c</sup>* |  | 1 | 1 | 2 |  |  |  | 3 | 1 | 8 |
| *Clinician-users; n<sup>c</sup>* | 2 | 3 | 1 | 2 |  | 1 | 1 | 1 | 1 | 12 |
| *Experts; n<sup>c</sup>* | 1 |  | 1 |  |  |  |  | 1 |  | 3 |
| *Adults only; n* | 19 | 7 | 1 | 10 | 2 | 3 | 5 | 11 | 7 | 65 |
| *Children only; n* |  |  | 1 | 1 |  | 2 |  | 3 | 3 | 10 |
| *Both adults and children; n* |  | 1 |  | 2 | 1 | 1 |  | 1 |  | 6 |
| *Not reported; n* |  | 1 | 1 |  |  |  |  |  |  | 2 |
| *Males/men only; n* |  |  |  |  |  |  |  | 1 | 2 | 3 |
| *Females/women only; n* | 1 |  |  |  | 1 | 1 |  |  | 2 | 5 |
| *Both or all sexes/genders; n* | 18 | 7 | 2 | 13 | 2 | 5 | 5 | 13 | 6 | 71 |
| *Not reported; n* |  | 2 | 1 |  |  |  |  | 1 |  | 4 |
| *Race/ethnicity reported; n* | 2 | 1 |  | 4 |  | 2 |  | 3 | 2 | 14 |
| *Race/ethnicity not reported; n* | 17 | 8 | 3 | 9 | 3 | 4 | 5 | 12 | 8 | 69 |
| **Number of sDHTs assessed** |  |  |  |  |  |  |  |  |  |  |
| *Range (min - max)* | 1 - 7 | 1 - 3 | 1 - 1 | 1 - 5 | 1 - 6 | 1 - 5 | 1 - 2 | 1 - 7 | 1 - 11 | 1 - 11 |

<sup>a</sup> 'Other' therapeutic area category contains studies with enrollment eligibility focused on anaphylaxis, muscular dystrophy, hemophilia, nocturnal enuresis, blood and marrow transplant, overweight/obesity, pregnancy, and non-specific hospitalized or chronic illness. One study recruited clinicians only (no end-users of the sDHT) and is included in this category.
<sup>b</sup> Studies reporting formative and summative evaluations are categorized as summative.
<sup>c</sup> Categories are not mutually-exclusive

### Sample characteristics

As shown in Table 3, the largest target populations were focused on aging and healthy participants (15 and 19 studies, respectively; 41% of all studies). Among the various diseases studied, neurology and cardiovascular were the most common therapeutic areas (13 and 9 studies, respectively; 50% of studies assessing non-healthy individuals). Supplementary Table 3 contains a list of conditions falling into each therapeutic area.

Almost all studies (82 of 83) captured data from targeted end-users; the remaining study captured data only from clinician-users [37]. Several studies captured data from multiple user groups; in total, 8 and 12 studies gathered data from carepartner-users and clinician-users, respectively. Three studies involved experts (not considered to be sDHT users); two of these described a formal heuristic evaluation [38,39] while the other described involving experts in design, biomedical engineering, computer science, and mHealth system production in the sDHTs design and formative testing process [40]. Finally, we noted substantial missing participant demographic data; age, sex/gender, and race/ethnicity were not reported in two, four, and 69 studies, respectively.

### sDHTs assessed in eligible studies

Across the 83 studies included in our review, a total of 164 different sDHTs were assessed (141 wearable and 23 ambient tools; see Table 4), ranging from 1 to 11 sDHTs within a single study. Ingestible and implantable sDHTs were in-scope, but none were identified in our literature search. A wide range of form factors (22 distinct categories) and wear locations (14 anatomical locations presented in five categories) were identified. Digital clinical measures of vital signs (n=76 sDHTs), physical activity (n=61 sDHTs), and mobility (n=35) were most prevalent. Supplementary Table 4 contains more comprehensive information regarding wear locations and health concepts captured by sDHTs.

**Table 4:**
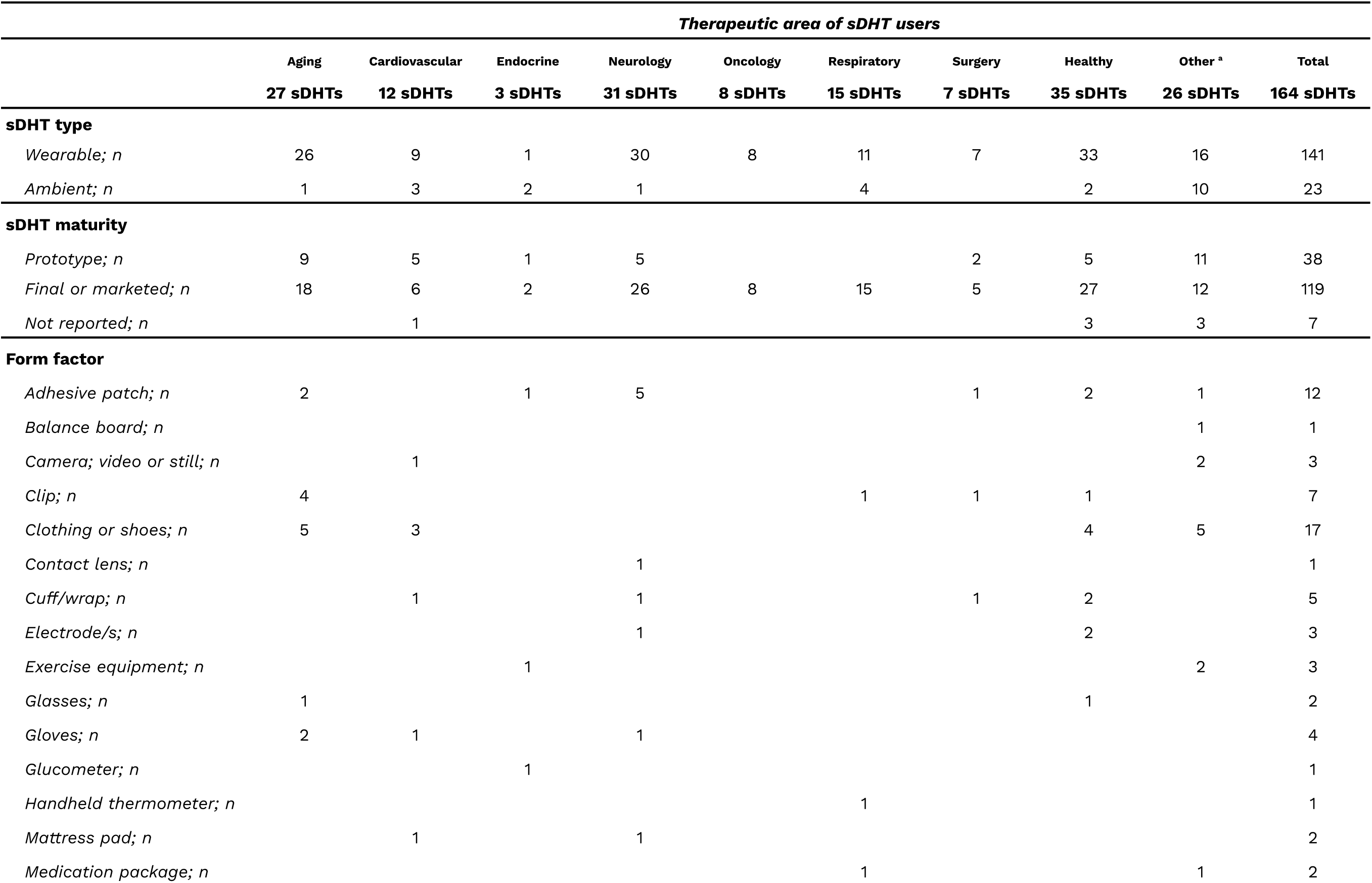
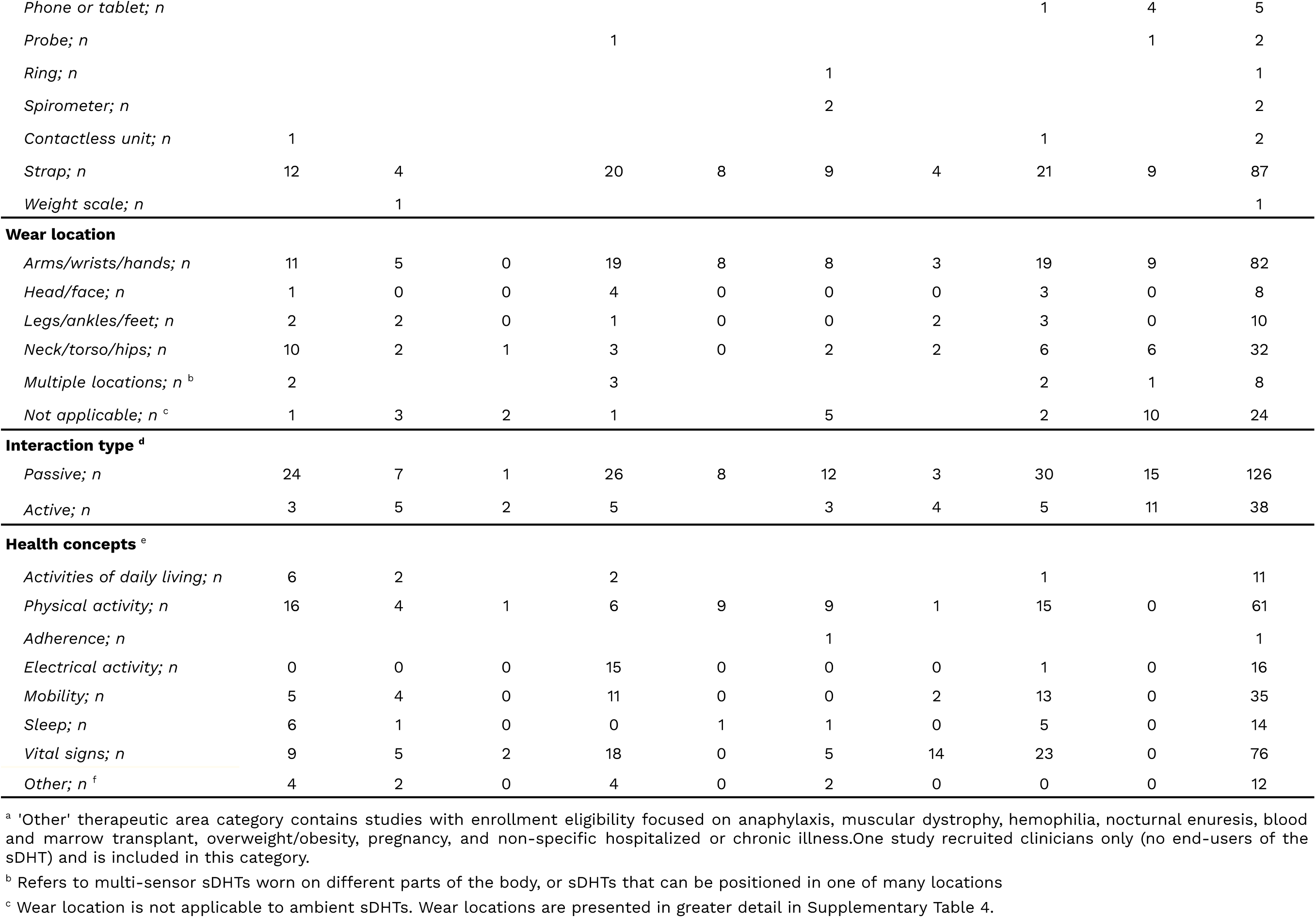
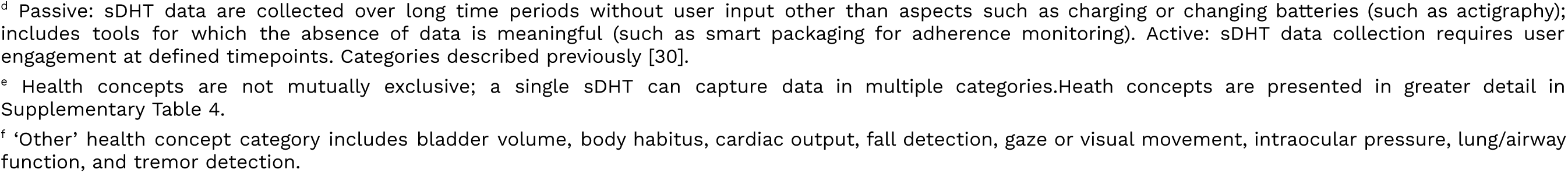
Sensor-based digital health technology descriptive information across therapeutic areas.

Most sDHTs (126 of 164) required only passive interaction by users, meaning that data were captured without user input other than basic tasks such as charging or changing batteries. The remaining 38 sDHTs required active engagement at specific times, such as completion of physical therapy [41], exercise [42], or blood glucose tests [39].

### Methodological approaches

As described in Table 5, most sDHTs (n=139) were evaluated in the actual environment in which they were intended to be used, while 25 were assessed in a simulated environment only. The vast majority were evaluated during actual use (n=148) rather than through “look and feel” approaches. Of particular interest, a variety of methods were used to evaluate usability and related concepts, including interviews (49 sDHTs), focus groups (29 sDHTs), direct or video observation (35 sDHTs), think-aloud (15 sDHTs), and heuristic analysis (2 sDHTs). Surveys were the most prevalent method for capturing usability data; 86 sDHTs were evaluated using referenced surveys while 81 were evaluated using surveys developed in-house by study investigators. Data for four sDHTs were captured using the sDHT itself; for example, instances of connectivity loss or data capture drops were recorded as use-errors and/or technical performance/product-errors [43,44].

**Table 5:**
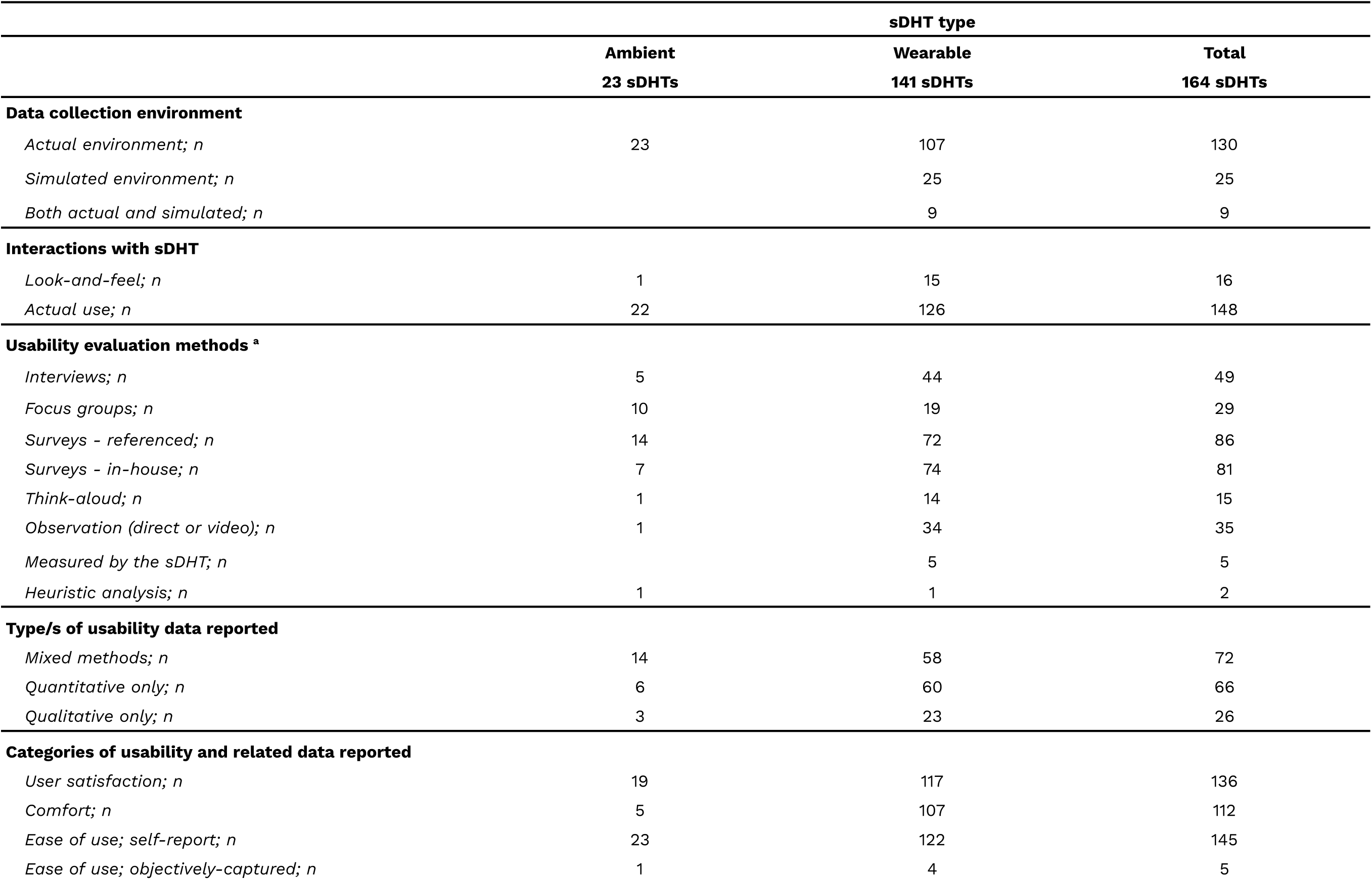
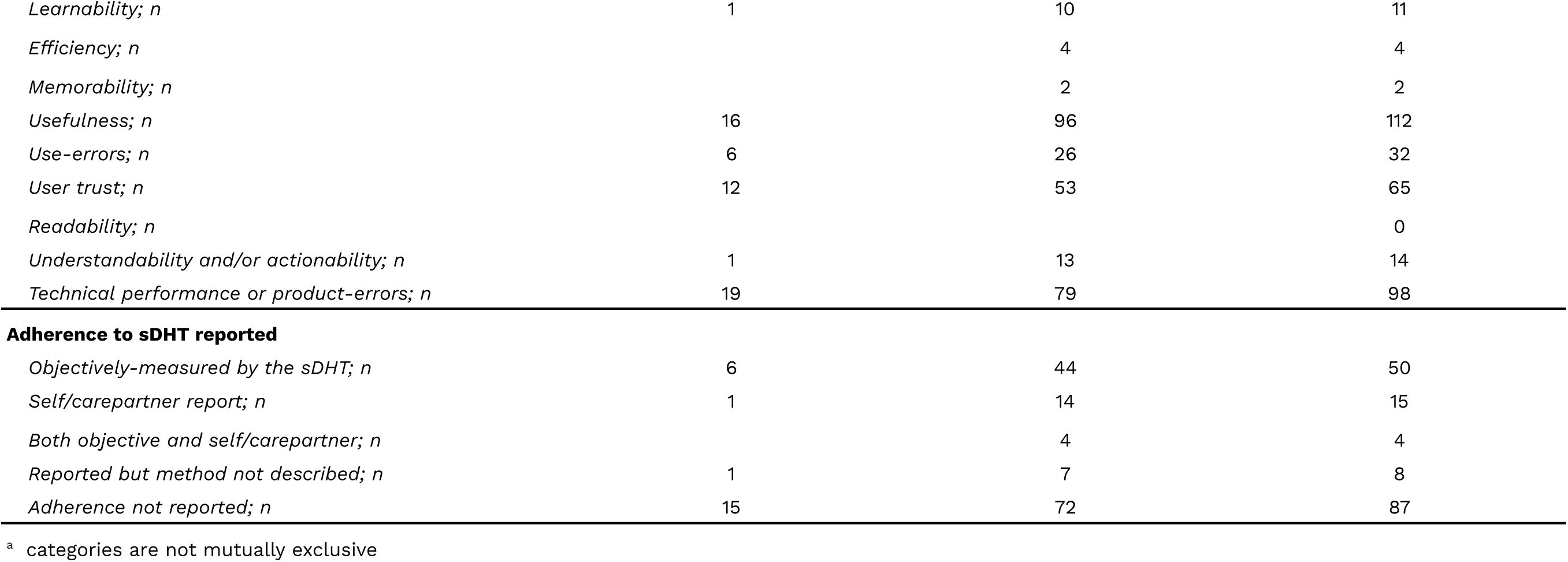
Methodological approaches to usability data collection.

|  | sDHT type |  |  |
| --- | --- | --- | --- |
|  | Ambient<br>23 sDHTs | Wearable<br>141 sDHTs | Total<br>164 sDHTs |
| **Data collection environment** |  |  |  |
| *Actual environment; n* | 23 | 107 | 130 |
| *Simulated environment; n* |  | 25 | 25 |
| *Both actual and simulated; n* |  | 9 | 9 |
| **Interactions with sDHT** |  |  |  |
| *Look-and-feel; n* | 1 | 15 | 16 |
| *Actual use; n* | 22 | 126 | 148 |
| **Usability evaluation methods <sup>a</sup>** |  |  |  |
| *Interviews; n* | 5 | 44 | 49 |
| *Focus groups; n* | 10 | 19 | 29 |
| *Surveys - referenced; n* | 14 | 72 | 86 |
| *Surveys - in-house; n* | 7 | 74 | 81 |
| *Think-aloud; n* | 1 | 14 | 15 |
| *Observation (direct or video); n* | 1 | 34 | 35 |
| *Measured by the sDHT; n* |  | 5 | 5 |
| *Heuristic analysis; n* | 1 | 1 | 2 |
| **Type/s of usability data reported** |  |  |  |
| *Mixed methods; n* | 14 | 58 | 72 |
| *Quantitative only; n* | 6 | 60 | 66 |
| *Qualitative only; n* | 3 | 23 | 26 |
| **Categories of usability and related data reported** |  |  |  |
| *User satisfaction; n* | 19 | 117 | 136 |
| *Comfort; n* | 5 | 107 | 112 |
| *Ease of use; self-report; n* | 23 | 122 | 145 |
| *Ease of use; objectively-captured; n* | 1 | 4 | 5 |
| *Learnability; n* | 1 | 10 | 11 |
| *Efficiency; n* |  | 4 | 4 |
| *Memorability; n* |  | 2 | 2 |
| *Usefulness; n* | 16 | 96 | 112 |
| *Use-errors; n* | 6 | 26 | 32 |
| *User trust; n* | 12 | 53 | 65 |
| *Readability; n* |  |  | 0 |
| *Understandability and/or actionability; n* | 1 | 13 | 14 |
| *Technical performance or product-errors; n* | 19 | 79 | 98 |
| **Adherence to sDHT reported** |  |  |  |
| *Objectively-measured by the sDHT; n* | 6 | 44 | 50 |
| *Self/carepartner report; n* | 1 | 14 | 15 |
| *Both objective and self/carepartner; n* |  | 4 | 4 |
| *Reported but method not described; n* | 1 | 7 | 8 |
| *Adherence not reported; n* | 15 | 72 | 87 |

<sup>a</sup> categories are not mutually exclusive

### Categories of usability-related data reported

User satisfaction was captured for the majority of sDHTs (n=136), often as a measure of acceptance/acceptability or user attitudes. Although overall ease-of-use was also commonly-reported, captured through either self-report or objective methods (n=145 and n=5, respectively), the related concepts of learnability, efficiency, and memorability were reported for only 11, 4, and 2 sDHTs, respectively. Technical performance and product-errors associated with malfunction were captured for 98 sDHTs, while use-errors were captured for only 32 sDHTs. Finally, although none of the studies in our review reported the readability of information presented to the user, 14 sDHTs were evaluated according to the extent to which users were able to understand the data or information presented to them (understandability) and/or the actions or tasks they should complete in response (actionability).

Finally, adherence (such as wear- or use-time) was reported for 77 sDHTs. Of these, 50 sDHTs captured adherence data objectively, adherence to 19 sDHTS was assessed through self- and/or carepartner-report, and the method was not described for eight sDHTs.

The complexity of the relationships in our dataset comparing usability evaluation methods with sDHT form factor, and comparing usability evaluation methods with the categories of usability-related data reported, are depicted in Figure 2 and Figure 3, respectively. For example, the width of each chord in Figure 2 is proportional to the number of sDHTs of the relevant form factor that were assessed using the linked method.

**Figure 2:**
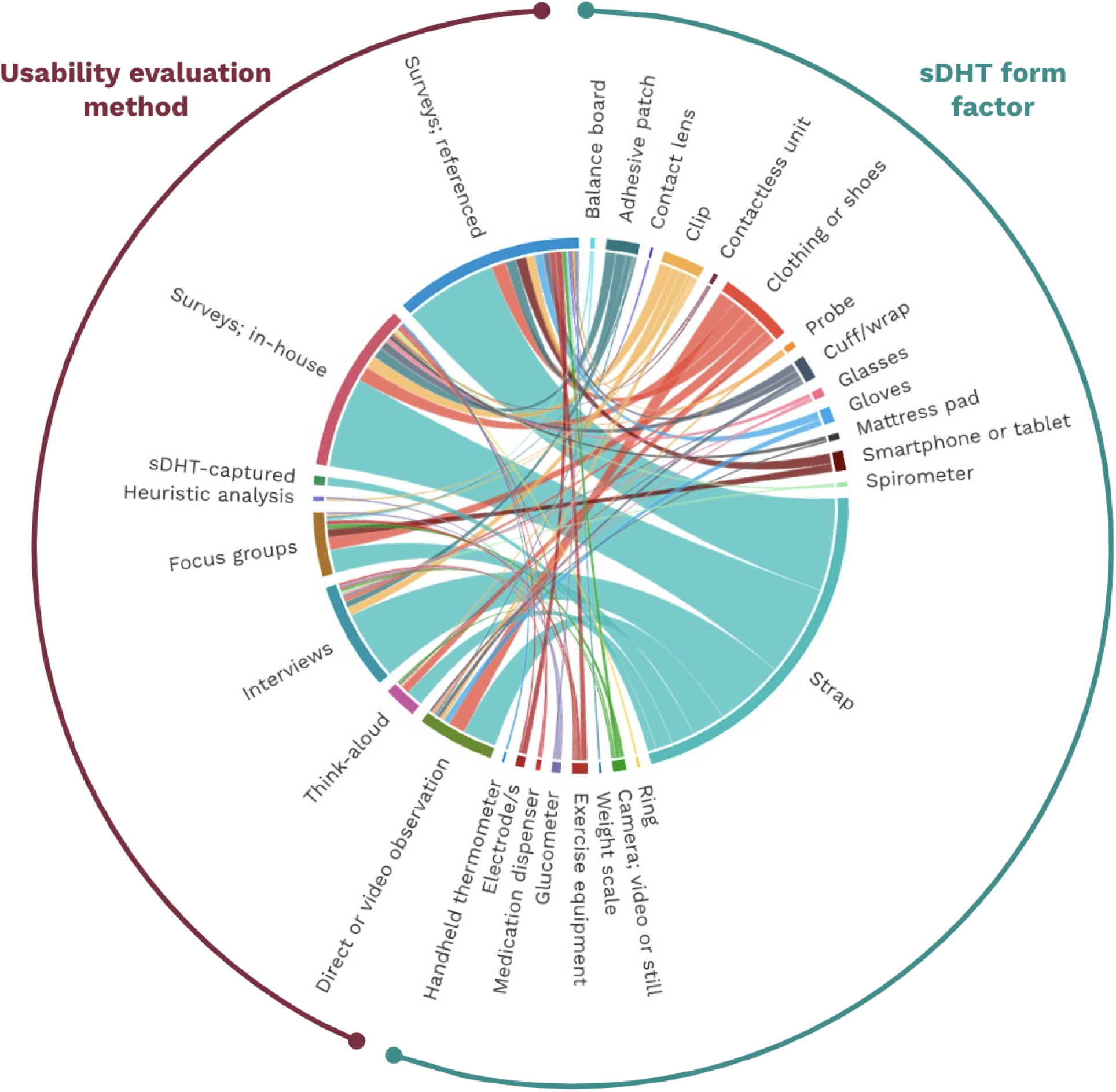
Chord diagram depicting the relationship between sDHT form factors and usability evaluation methods

**Figure 3:**
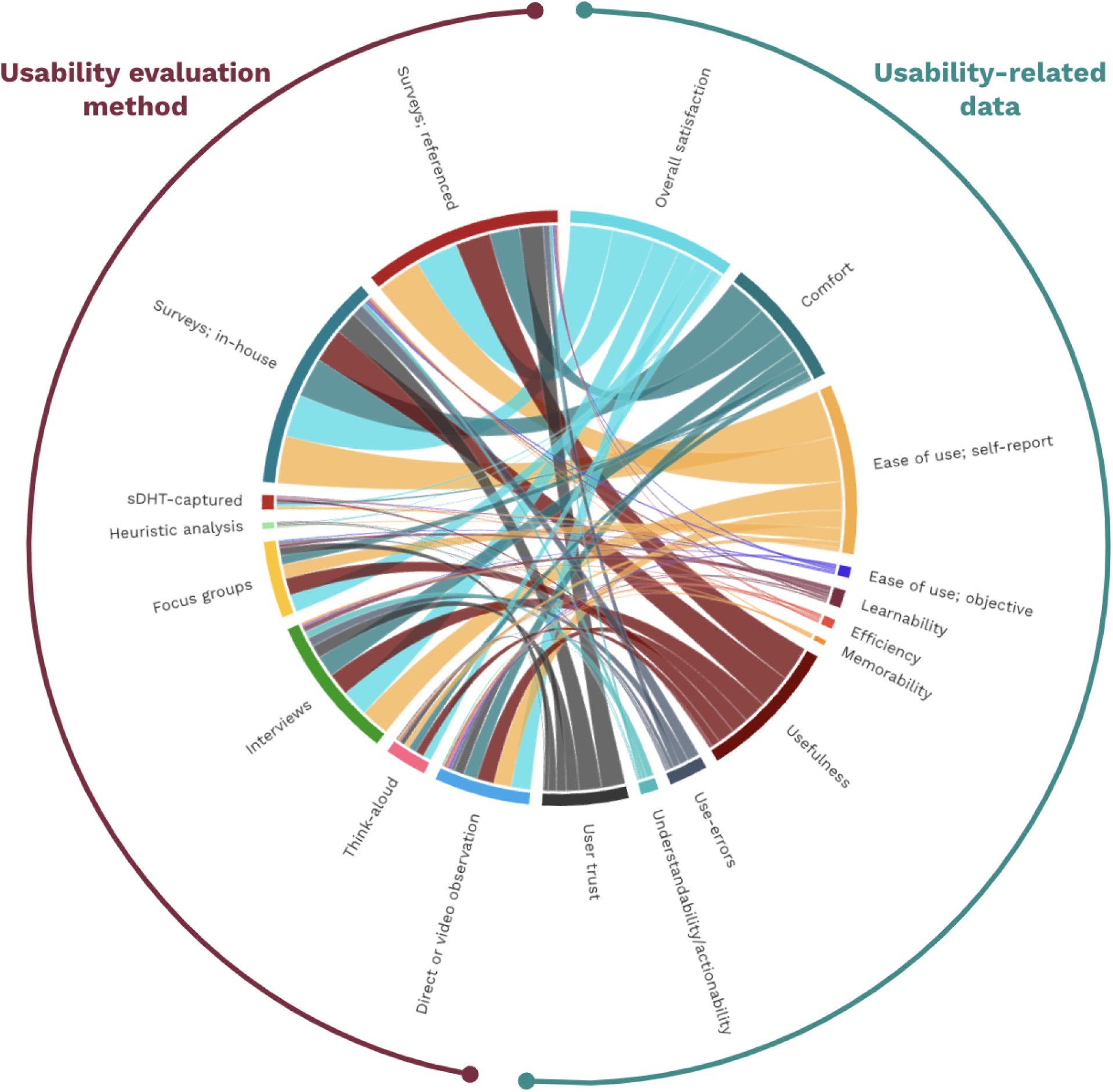
Chord diagram depicting the relationship between usability evaluation methods and categories of usability-related data reported

Supplementary Tables 5-7 present the data shown in Tables 3-5 for the subset of 55 studies reporting the results of summative evaluations, while Supplementary Tables 8-10 present these data for the subset of 28 studies reporting formative evaluations.

## Discussion

### Principal findings

This manuscript represents the first systematic scoping review reporting the methodological approaches adopted during usability-related studies specifically focused on sDHTs. We identified 83 formative and summative studies published over the decade from 2003-2023 which evaluated human factors, human-centered design, and/or usability for 164 ambient and wearable tools. Most studies (67 of 83; 81%) recruited non-healthy individuals, thereby providing informative data regarding sDHT usability across many diseases as well as conditions such as ageing and pregnancy. Most sDHTs were evaluated in the intended use environment, with multiple facets of usability-related data captured via a range of mixed method approaches including heuristic analysis, surveys, observation, think-aloud, focus groups, interviews, and use and/or technical performance/product-errors captured by the sDHT itself such as instances of connectivity loss.

This review highlights four notable gaps that warrant attention as the field advances. Firstly, the breadth and scope of usability and related data were fairly simplistic, relying largely on surveys capturing user satisfaction and ease of use (each captured for >80% of sDHTs) with limited reporting of sDHT use-errors, learnability, efficiency, or memorability. The extent to which users understood the clinical data or other information presented to them (understandability) and the actions or tasks they should complete in response (actionability) was assessed for only 9% of sDHTs. For the use of sDHTs in clinical care settings, it is imperative that users understand whether and how to react to clinical data [45], and thus the lack of focus on understandability and actionability is concerning and could be due to the early-stage nature of sDHTs in clinical practice. In the context of clinical research, however, sharing sDHT data with participants in real time has the potential to introduce bias and affect user behavior, thereby posing a risk of yielding inaccurate results [4]. Secondly, only 22% of studies considered users other than end-users (patients/participants), such as carepartners and clinicians, who play crucial roles in sDHT implementation and therefore the quality of data captured [46]. Especially in populations where carepartners play a key role in sDHT implementation (for example, children, elderly individuals, those with language barriers, and those with disabilities), understanding usability from the carepartner perspective is vital. Thirdly, we found that only 30% of summative studies (referred to by the FDA as ‘human factors validation studies’; [23]) provided a rationale for the sample size, with or without a power calculation. An understanding of key study design considerations including sample size are important for evaluating the robustness of study conclusions. Finally, as has been noted previously [30,47], we observed a deficiency in reporting basic sample demographics, with studies typically providing information on age and sex/gender but neglecting to include details on the race/ethnicity of participants. Inadequate reporting of descriptive data including sociodemographics precludes a complete understanding of generalizability, potentially leading to the need to repeat studies while contributing to disparities and biases in clinical research [48].

While several systematic reviews have focused on understanding and quantifying usability of digital health products for specific applications [49–53], few have focused on evaluating methodological approaches rather than study outcomes. Of those that have, most have focused on digital health technologies such as electronic medical records systems [54] and mobile clinical decision support tools [55] that are not used for remote data capture. In 2023, Maqbool and Herold [8] published a systematic review of usability evaluations describing a broad suite of over 1000 digital health tools consisting mostly of mHealth applications and including a subset of 20 products approximately aligned to our definition of sDHT, including fitness/activity trackers, digital sphygmomanometers, and wearable fall risk assessment systems. Compared to our study, Maqbool and Herold found relatively increased rates of clinician and carepartner participation, and reporting of learnability, efficiency, and memorability. Such differences emphasize substantial variability in usability study methodology across sub-categories of digital health technologies, as well as differences in definitions and terminology of the concepts reported, underscoring the need for a common evaluation framework.

### Strengths and limitations

Strengths of our review include the robust approach taken to testing our search terms, including a careful assessment against a list of target papers identified *a priori* to ensure that we were capturing appropriate literature. This process was intended to not only ensure the inclusivity of relevant literature but also the reliability of our findings to help provide a foundation for subsequent reviews and meta-analyses. In-depth data extraction across many domains allowed for a thorough comparative analysis of the identified studies. The decision to focus on studies published within the last decade (2013-2023) was also carefully considered, as it encompasses the recent surge in studies reporting sDHT implementation. While sDHTs have a lengthy history prior to 2013, this temporal scope ensures that our findings reflect contemporary developments and trends, offering insights into the current state of sDHT implementation.

A number of limitations are acknowledged. Firstly, we limited our search to the peer-reviewed literature. We acknowledge that many usability studies undertaken by technology manufacturers may be published in the gray literature; however, our ultimate goal is to use the findings of our review to guide development of a framework representing best practices, and therefore the peer-review process was used as an indicator of methodological rigor and reporting quality. Secondly, terminology in the field of digital medicine is still evolving and investigators use many different terms to describe sDHTs; by incorporating 25 descriptive keywords in Layer C of our search terms (see Supplementary Table 1), we found it necessary to rely on MeSH terms developed by the National Library of Medicine [28] as a means of limiting our literature search to a feasible number of publications. As a consequence, we were limited to conducting our search in PubMed as this is the clinical research database for the National Library of Medicine. While MeSH terms are widely accepted and systematically applied, their specificity may have excluded relevant studies using different terminology potentially resulting in unintentional omissions. Our hope is that as the field matures, terminology will become harmonized and sDHT-specific indexing will support the identification of studies adopting these technologies. Finally, only 40% of publications were screened for eligibility by multiple investigators. This approach to study identification, which has been described and adopted previously [29,30], allowed us to screen a greater number of papers which was necessary given the lack of systematic indexing. The high agreement levels between investigators suggest that our quality-control approach maintained a robust screening process, despite part of the work being conducted by a single investigator.

### Conclusions and future directions

Based on our findings, we suggest four actionable recommendations that will help to advance the implementation of sensor-based digital measurement tools in both clinical and research settings. Firstly, we encourage investigators to adopt in-depth assessment and reporting of usability data beyond user satisfaction and ease of use. In particular, it is valuable to understand use-errors alongside technical errors, and it is critical to evaluate the extent to which users understand the clinical data and information presented to them and the appropriate tasks to undertake in response, if applicable. Secondly, it is essential to embrace diversity of users in all respects, including evaluation of usability across multiple user groups including carepartners and clinicians, as well as ensuring that the participating users are generalizable to the intended use population in terms of sociodemographics, social determinants of health, and other characteristics. Thirdly, rigorous study design is key. Usability is a heterogeneous concept, and it is often beneficial to evaluate usability alongside other objectives such as analytical or clinical validation; thus, we do not advocate a particular study design or set of study outcome measures. We do, however, believe that careful consideration of usability evaluation criteria, study sample sizes, and predetermined thresholds of success are critical for making go/no-go decisions as to whether a particular sDHT is sufficiently usable for implementation in a particular context of use. Lastly, we recommend adhering to reporting and publication checklists such as Annex B in ISO 9241-11:2018 [56] and/or EVIDENCE [57], the latter of which describes optimal reporting requirements of studies evaluating several aspects of sDHT quality including usability assessments. Ensuring consistency in reporting will enable meaningful comparisons between studies, facilitate better assessments of findings, and enhance the accurate interpretation of results and limitations across studies.

Our long-term goal is to develop and disseminate an evidence-driven framework for evaluating sDHTs as being fit-for-purpose from a usability perspective, informed in part from the findings of this review. By developing such a framework, we endeavor to contribute to the ongoing discourse surrounding sDHTs, ultimately paving the way for the development of safe and effective tools that lead to a more inclusive and patient-centric healthcare ecosystem poised to improve clinical trials and clinical practice.

## Data Availability

All data produced in the present study are available upon reasonable request to the authors

## Acknowledgements

The authors wish to acknowledge Bethanie McCrary and Danielle Stefko for assistance with project management; Katerina Djambazova for assistance with data extraction; and Jennifer Goldsack for providing feedback on the manuscript. Chord diagrams were created with flourish.studio online software.

All authors contributed to study design, data interpretation, and manuscript preparation. The literature search, literature screening, and data extraction were undertaken by JC, SM, JPB, and Katerina Djambazova. This work was undertaken within the Digital Health Measurement Collaborative Community (<u>DATAcc</u>), hosted by the Digital Medicine Society (<u>DiMe</u>).

## Abbreviations

DATAcc: Digital Health Measurement Collaborative Community
DiMe: Digital Medicine Society
FDA: Food and Drug Administration (United States)
ISO: International Organization for Standardization
MeSH: Medical Subject Heading
MHRA: Medicines and Healthcare products Regulatory Agency (United Kingdom)
NMPA: National Medical Products Administration (China)
PICO: Patients/participants; intervention; comparator; outcomes
sDHT: Sensor-based digital health technology

