## Supplementary Table for "A systematic scoping review of studies describing human factors, human-centered design, and usability of sensor-based digital health technologies"

### Supplementary Table 1: PubMed search terms

| Layer | Search terms | Purpose | Papers identified; *n* |
|---|---|---|---|
| A | Humans[mesh] | Limits the search to studies of human participants | 7,245,671 |
| AND | | | |
| B | Wearable electronic devices[mesh] OR<br>Wireless technology[mesh] OR<br>Remote sensing technology[mesh] OR<br>Digital technology[mesh] | MeSH terms relevant to digital health technologies | 20,896 |
| AND | | | |
| C | BioMeT[tiab] OR<br>Biometric monitor*[tiab] OR<br>Digital health[tiab] OR<br>Digital med*[tiab] OR<br>Digital sens*[tiab] OR<br>Biosensor[tiab] OR<br>Transducer[tiab] OR<br>Ingest*[tiab] OR<br>Implant*[tiab] OR<br>Wear*[tiab] OR<br>Worn[tiab] OR<br>Nearable[tiab] OR<br>Tracker[tiab] OR<br>Smart*[tiab] OR<br>Mobile[tiab] OR<br>Remote[tiab] OR<br>Home-based[tiab] OR<br>In-home[tiab] OR<br>Portable[tiab] OR<br>Ambulatory[tiab] OR<br>Wireless[tiab] OR<br>Connected[tiab] OR<br>Wifi[tiab] OR<br>Bluetooth[tiab] OR<br>Cloud[tiab] | Keywords relevant to digital health technologies | 697,121 |
| AND | | | |
| D | Usab*[tiab] OR | Keywords relevant to human-centered design, | 47,375 |

| | | | |
|---|---|---|---|
| | Ergonomic*[tiab] OR<br>Human-factor*[tiab] OR<br>User-cent*[tiab] OR<br>Human-cent*[tiab] OR<br>Wearability[tiab] OR<br>Summative[tiab] OR<br>Formative[tiab] OR<br>User-experience*[tiab] | human factors, usability, and related concepts | |
| NOT | | | |
| E | Case Reports[pt] OR<br>Clinical Trial Protocol[pt] OR<br>Comment[pt] OR<br>Editorial[pt] OR<br>English Abstract[pt] OR<br>Letter[pt] OR<br>Meta-Analysis[pt] OR<br>Review[pt] OR<br>Systematic Review[pt] | Excludes publication types other than clinical research studies | 3,140,569 |
| AND | | | |
| F | 2013/01/01:2023/05/30[pdat] | Limits the search to a 10-year date range | 13,190,723 |

**Complete search string returning 442 results as of June 1st, 2023:**
(((((Humans[mesh]) AND (Wearable electronic devices[mesh] OR Wireless technology[mesh] OR Remote sensing technology[mesh] OR Digital technology[mesh])) AND (BioMeT[tiab] OR Biometric monitor*[tiab] OR Digital health[tiab] OR Digital med*[tiab] OR Digital sens*[tiab] OR Biosensor[tiab] OR Transducer[tiab] OR Ingest*[tiab] OR Implant*[tiab] OR Wear*[tiab] OR Worn[tiab] OR Nearable[tiab] OR Tracker[tiab] OR Smart*[tiab] OR Mobile[tiab] OR Remote[tiab] OR Home-based[tiab] OR In-home[tiab] OR Portable[tiab] OR Ambulatory[tiab] OR Wireless[tiab] OR Connected[tiab] OR Wifi[tiab] OR Bluetooth[tiab] OR Cloud[tiab])) AND (Usab*[tiab] OR Ergonomic*[tiab] OR Human-factor*[tiab] OR User-cent*[tiab] OR Human-cent*[tiab] OR Wearability[tiab] OR Summative[tiab] OR Formative[tiab] OR User-experience*[tiab])) NOT (Case Reports[pt] OR Clinical Trial Protocol[pt] OR Comment[pt] OR Editorial[pt] OR English Abstract[pt] OR Letter[pt] OR Meta-Analysis[pt] OR Review[pt] OR Systematic Review[pt])) AND (2013/01/01:2023/05/30[pdat])

Search tags in square brackets are as follows: mesh = Medical subject headings; tiab = Title and abstract*; pt = Publication type; pdat = Publication date.
* Keywords are searched within the citation title, collection title, abstract, and author keywords.

### Supplementary Table 2: All included studies

| Title | Authors | Citation | DOI | Pubmed ID |
|---|---|---|---|---|
| Continuous Monitoring of Vital Signs in the General Ward Using Wearable Devices: Randomized Controlled Trial | Weenk M, Bredie SJ, Koeneman M, Hesselink G, van Goor H, van de Belt TH. | J Med Internet Res. 2020 Jun 10;22(6):e15471. doi: 10.2196/15471. | 10.2196/15471 | 32519972 |
| Remote Health Monitoring for Older Adults and Those with Heart Failure: Adherence and System Usability | Evans J, Papadopoulos A, Silvers CT, Charness N, Boot WR, Schlachta-Fairchild L, Crump C, Martinez M, Ent CB. | Telemed J E Health. 2016 Jun;22(6):480-8. doi: 10.1089/tmj.2015.0140. Epub 2015 Nov 5. | 10.1089/tmj.2015.0140 | 26540369 |
| The Use of a Smartphone App and an Activity Tracker to Promote Physical Activity in the Management of Chronic Obstructive Pulmonary Disease: Randomized Controlled Feasibility Study | Bentley CL, Powell L, Potter S, Parker J, Mountain GA, Bartlett YK, Farwer J, O'Connor C, Burns J, Cresswell RL, Dunn HD, Hawley MS. | JMIR Mhealth Uhealth. 2020 Jun 3;8(6):e16203. doi: 10.2196/16203. | 10.2196/16203 | 32490838 |
| User experiences and perceptions of health wearables: an exploratory study in Cambodia | Liverani M, Ir P, Wiseman V, Perel P. | Glob Health Res Policy. 2021 Sep 23;6(1):33. doi: 10.1186/s41256-021-00221-3. | 10.1186/s41256-021-00221-3 | 34556184 |
| Signal quality and patient experience with wearable devices for epilepsy management | Nasseri M, Nurse E, Glasstetter M, Böttcher S, Gregg NM, Laks Nandakumar A, Joseph B, Pal Attia T, Viana PF, Bruno E, Biondi A, Cook M, Worrell GA, Schulze-Bonhage A, Dümpelmann M, Freestone DR, Richardson MP, Brinkmann BH. | Epilepsia. 2020 Nov;61 Suppl 1:S25-S35. doi: 10.1111/epi.16527. Epub 2020 Jun 4. | 10.1111/epi.16527 | 32497269 |
| Digital technology for anaphylaxis management impact on patient behaviour: A randomized clinical trial | Sala-Cunill A, Luengo O, Curran A, Moreno N, Labrador-Horrillo M, Guilarte M, Gonzalez-Medina M, Galvan-Blasco P, Cardona V. | Allergy. 2021 May;76(5):1507-1516. doi: 10.1111/all.14626. Epub 2020 Nov 4. | 10.1111/all.14626 | 33043475 |
| Mobile Health Technology for Pediatric Symptom Monitoring: A Feasibility Study | Vaughn J, Gollarahalli S, Shaw RJ, Docherty S, Yang Q, Malhotra C, Summers-Goeckerman E, Shah N. | Nurs Res. 2020 Mar/Apr;69(2):142-148. doi: 10.1097/NNR.0000000000000403. | 10.1097/NNR.0000000000000403 | 31972852 |
| Wearable gait analysis systems: ready to be used by medical practitioners in geriatric wards? | Ollenschläger M, Kluge F, Müller-Schulz M, Püllen R, Möller C, Klucken J, Eskofier BM. | Eur Geriatr Med. 2022 Aug;13(4):817-824. doi: 10.1007/s41999-022-00629-1. Epub 2022 Mar 3. | 10.1007/s41999-022-00629-1 | 35243600 |
| Predictors of engagement with remote sensing technologies for symptom measurement in Major Depressive Disorder | Matcham F, Carr E, White KM, Leightley D, Lamers F, Siddi S, Annas P, de Girolamo G, Haro JM, Horsfall M, Ivan A, Lavelle G, Li Q, Lombardini F, Mohr DC, Narayan VA, Penninx BWHJ, Oetzmann C, Coromina M, Simblett SK, Weyer J, Wykes T, Zorbas S, Brasen JC, Myin-Germeys I, Conde P, Dobson RJB, Folarin AA, Ranjan Y, Rashid | J Affect Disord. 2022 Aug 1;310:106-115. doi: 10.1016/j.jad.2022.05.005. Epub 2022 May 5. | 10.1016/j.jad.2022.05.005 | 35525507 |

| | | | | |
|---|---|---|---|---|
| | Z, Cummins N, Dineley J, Vairavan S, Hotopf M; RADAR-CNS consortium. | | | |
| Patients self-mastery of wearable devices for seizure detection: A direct user-experience | Bruno E, Biondi A, Thorpe S, Richardson MP; RADAR-CNS Consortium. | Seizure. 2020 Oct;81:236-240. doi: 10.1016/j.seizure.2020.08.023. Epub 2020 Aug 23. | 10.1016/j.seizure.2020.08.023 | 32889512 |
| Wearable Inertial Sensor System Towards Daily Human Kinematic Gait Analysis: Benchmarking Analysis to MVN BIOMECH | Figueiredo J, Carvalho SP, Vilas-Boas JP, Gonçalves LM, Moreno JC, Santos CP. | Sensors (Basel). 2020 Apr 12;20(8):2185. doi: 10.3390/s20082185. | 10.3390/s20082185 | 32290636 |
| Feasibility and Acceptability of Wearable Sleep Electroencephalogram Device Use in Adolescents: Observational Study | Lunsford-Avery JR, Keller C, Kollins SH, Krystal AD, Jackson L, Engelhard MM. | JMIR Mhealth Uhealth. 2020 Oct 1;8(10):e20590. doi: 10.2196/20590. | 10.2196/20590 | 33001035 |
| A Smart Wearable Sensor System for Counter-Fighting Overweight in Teenagers | Standoli CE, Guarneri MR, Perego P, Mazzola M, Mazzola A, Andreoni G. | Sensors (Basel). 2016 Aug 10;16(8):1220. doi: 10.3390/s16081220. | 10.3390/s16081220 | 27517929 |
| Tolerability and Functionality of a Wireless 24-Hour Ocular Telemetry Sensor in African American Glaucoma Patients | Marando CM, Mansouri K, Kahook MY, Seibold LK. | J Glaucoma. 2019 Feb;28(2):119-124. doi: 10.1097/IJG.0000000000001141. | 10.1097/IJG.0000000000001141 | 30688840 |
| Feasibility and Usability Aspects of Continuous Remote Monitoring of Health Status in Palliative Cancer Patients Using Wearables | Pavic M, Klaas V, Theile G, Kraft J, Tröster G, Guckenberger M. | Oncology. 2020;98(6):386-395. doi: 10.1159/000501433. Epub 2019 Jul 23. | 10.1159/000501433 | 31336377 |
| Development of User-Friendly Wearable Electronic Textiles for Healthcare Applications | Yang K, Meadmore K, Freeman C, Grabham N, Hughes AM, Wei Y, Torah R, Glanc-Gostkiewicz M, Beeby S, Tudor J. | Sensors (Basel). 2018 Jul 25;18(8):2410. doi: 10.3390/s18082410. | 10.3390/s18082410 | 30044382 |
| Continuous Noninvasive Remote Automated Blood Pressure Monitoring With Novel Wearable Technology: A Preliminary Validation Study | McGillion MH, Dvirnik N, Yang S, Belley-Côté E, Lamy A, Whitlock R, Marcucci M, Borges FK, Duceppe E, Ouellette C, Bird M, Carroll SL, Conen D, Tarride JE, Harsha P, Scott T, Good A, Gregus K, Sanchez K, Benoit P, Owen J, Harvey V, Peter E, Petch J, Vincent J, Graham M, Devereaux PJ. | JMIR Mhealth Uhealth. 2022 Feb 28;10(2):e24916. doi: 10.2196/24916. | 10.2196/24916 | 34876396 |
| Physical Activity Monitoring and Acceptance of a Commercial Activity Tracker in Adult Patients with Haemophilia | Carrasco JJ, Pérez-Alenda S, Casaña J, Soria-Olivas E, Bonanad S, Querol F. | Int J Environ Res Public Health. 2019 Oct 12;16(20):3851. doi: 10.3390/ijerph16203851. | 10.3390/ijerph16203851 | 31614706 |
| Wearable Sensor-Based Exercise Biofeedback for Orthopaedic Rehabilitation: A Mixed Methods User Evaluation of a Prototype System | Argent R, Slevin P, Bevilacqua A, Neligan M, Daly A, Caulfield B. | Sensors (Basel). 2019 Jan 21;19(2):432. doi: 10.3390/s19020432. | 10.3390/s19020432 | 30669657 |
| A Mixed-Methods Study of Users' Journey Mapping Experience and Acceptance of Telehealthcare Technology in Taiwan | Tsung-Yin O, Chih-Young H, Che-Wei L. | Telemed J E Health. 2019 Nov;25(11):1057-1070. doi: 10.1089/tmj.2018.0155. Epub 2019 Jan 29. | 10.1089/tmj.2018.0155 | 30694730 |
| Patients' experience of wearing | Simblett SK, Biondi A, Bruno E, Ballard D, | Epilepsy Behav. 2020 Jan;102:106717. doi: | 10.1016/j.yebeh. | 31785481 |

| | | | | |
|---|---|---|---|---|
| multimodal sensor devices intended to detect epileptic seizures: A qualitative analysis | Stoneman A, Lees S, Richardson MP, Wykes T; RADAR-CNS consortium. | 10.1016/j.yebeh.2019.106717. Epub 2019 Nov 27. | 2019.106717 | |
| Feasibility of remote digital monitoring using wireless Bluetooth monitors, the Smart Angel™ app and an original web platform for patients following outpatient surgery: a prospective observational pilot study | Chevallier T, Buzancais G, Occean BV, Rataboul P, Boisson C, Simon N, Lannelongue A, Chaniaud N, Gricourt Y, Lefrant JY, Cuvillon P. | BMC Anesthesiol. 2020 Oct 8;20(1):259. doi: 10.1186/s12871-020-01178-5. | 10.1186/s12871-020-01178-5 | 33032541 |
| Body-Worn Sensors for Parkinson's disease: A qualitative approach with patients and healthcare professionals | Virbel-Fleischman C, Rétory Y, Hardy S, Huiban C, Corvol JC, Grabli D. | PLoS One. 2022 May 5;17(5):e0265438. doi: 10.1371/journal.pone.0265438. eCollection 2022. | 10.1371/journal.pone.0265438 | 35511812 |
| User-based evaluation of applicability and usability of a wearable accelerometer device for detecting bilateral tonic-clonic seizures: A field study | Meritam P, Ryvlin P, Beniczky S. | Epilepsia. 2018 Jun;59 Suppl 1:48-52. doi: 10.1111/epi.14051. | 10.1111/epi.14051 | 29873828 |
| Safety and usability of wearable accelerometers for stroke detection the STROKE ALARM PRO 1 study | Esbjörnsson M, Ullberg T. | J Stroke Cerebrovasc Dis. 2022 Nov;31(11):106762. doi: 10.1016/j.jstrokecerebrovasdis.2022.106762. Epub 2022 Sep 15. | 10.1016/j.jstrokecerebrovasdis.2022.106762 | 36115106 |
| Design of a Low-Cost, Wearable Device for Kinematic Analysis in Physical Therapy Settings | Hua A, Johnson N, Quinton J, Chaudhary P, Buchner D, Hernandez ME. | Methods Inf Med. 2020 Feb;59(1):41-47. doi: 10.1055/s-0040-1710380. Epub 2020 Jun 14. | 10.1055/s-0040-1710380 | 32535880 |
| Usability, Acceptability, and Satisfaction of a Wearable Activity Tracker in Older Adults: Observational Study in a Real-Life Context in Northern Portugal | Domingos C, Costa P, Santos NC, Pêgo JM. | J Med Internet Res. 2022 Jan 26;24(1):e26652. doi: 10.2196/26652. | 10.2196/26652 | 35080503 |
| Recommendations for Identifying Valid Wear for Consumer-Level Wrist-Worn Activity Trackers and Acceptability of Extended Device Deployment in Children | Wing D, Godino JG, Baker FC, Yang R, Chevance G, Thompson WK, Reuter C, Bartsch H, Wilbur A, Straub LK, Castro N, Higgins M, Colrain IM, de Zambotti M, Wade NE, Lisdahl KM, Squeglia LM, Ortigara J, Fuemmeler B, Patrick K, Mason MJ, Tapert SF, Bagot KS. | Sensors (Basel). 2022 Nov 26;22(23):9189. doi: 10.3390/s22239189. | 10.3390/s22239189 | 36501894 |
| Lumbatex: A Wearable Monitoring System Based on Inertial Sensors to Measure and Control the Lumbar Spine Motion | Cortell-Tormo JM, Garcia-Jaen M, Ruiz-Fernandez D, Fuster-Lloret V. | IEEE Trans Neural Syst Rehabil Eng. 2019 Aug;27(8):1644-1653. doi: 10.1109/TNSRE.2019.2927083. Epub 2019 Jul 5. | 10.1109/TNSRE.2019.2927083 | 31283484 |
| Comparing the Usability and Acceptability of Wearable Sensors Among Older Irish Adults in a Real-World Context: Observational Study | Keogh A, Dorn JF, Walsh L, Calvo F, Caulfield B. | JMIR Mhealth Uhealth. 2020 Apr 20;8(4):e15704. doi: 10.2196/15704. | 10.2196/15704 | 32310149 |
| Usability of affordable feedback-based | Hamilton C, Lovarini M, van den Berg M, | Disabil Rehabil. 2022 | 10.1080/096382 | 33645384 |

| | | | | |
|---|---|---|---|---|
| technologies to improve mobility and physical activity in rehabilitation: a mixed methods study | McCluskey A, Hassett L. | Jul;44(15):4029-4038. doi: 10.1080/09638288.2021.1884904. Epub 2021 Feb 28. | 88.2021.188490 4 | |
| Measure It Super Simple (MISS) activity tracker: (re)design of a user-friendly interface and evaluation of experiences in daily life | Ummels D, Braun S, Stevens A, Beekman E, Beurskens A. | Disabil Rehabil Assist Technol. 2022 Oct;17(7):767-777. doi: 10.1080/17483107.2020.1815089. Epub 2020 Sep 24. | 10.1080/1748310 7.2020.1815089 | 32970493 |
| Older Women's Experiences of a Community-Led Walking Programme Using Activity Trackers | O'Brien J, Mason A, Cassarino M, Chan J, Setti A. | Int J Environ Res Public Health. 2021 Sep 17;18(18):9818. doi: 10.3390/ijerph18189818. | 10.3390/ijerph1 8189818 | 34574741 |
| Usability and Interoperability in Wireless Sensor Networks for Patient Telemonitoring in Chronic Disease Management | Jiménez-Fernández S, de Toledo P, del Pozo F. | IEEE Trans Biomed Eng. 2013 Dec;60(12):3331-9. doi: 10.1109/TBME.2013.2280967. Epub 2013 Sep 5. | 10.1109/TBME.2 013.2280967 | 24021636 |
| Wearability Testing of Ambulatory Vital Sign Monitoring Devices: Prospective Observational Cohort Study | Areia C, Young L, Vollam S, Ede J, Santos M, Tarassenko L, Watkinson P. | JMIR Mhealth Uhealth. 2020 Dec 16;8(12):e20214. doi: 10.2196/20214. | 10.2196/20214 | 33325827 |
| Day and night comfort and stability on the body of four wearable devices for seizure detection: A direct user-experience | Bruno E, Biondi A, Böttcher S, Lees S, Schulze-Bonhage A, Richardson MP; RADAR-CNS Consortium. | Epilepsy Behav. 2020 Nov;112:107478. doi: 10.1016/j.yebeh.2020.107478. Epub 2020 Sep 28. | 10.1016/j.yebeh. 2020.107478 | 33181896 |
| Assessing the Clinical Utility of a Wearable Device for Physiological Monitoring of Heart Rate Variability in Military Service Members with Traumatic Brain Injury | Uomoto JM, Skopp N, Jenkins-Guarnieri M, Reini J, Thomas D, Adams RJ, Tsui M, Miller SR, Scott BR, Pasquina PF. | Telemed J E Health. 2022 Oct;28(10):1496-1504. doi: 10.1089/tmj.2021.0627. Epub 2022 Feb 28. | 10.1089/tmj.202 1.0627 | 35231193 |
| NeuroSuitUp: System Architecture and Validation of a Motor Rehabilitation Wearable Robotics and Serious Game Platform | Mitsopoulos K, Fiska V, Tagaras K, Papias A, Antoniou P, Nizamis K, Kasimis K, Sarra PD, Mylopoulou D, Savvidis T, Praftsiotis A, Arvanitidis A, Lyssas G, Chasapis K, Moraitopoulos A, Astaras A, Bamidis PD, Athanasiou A. | Sensors (Basel). 2023 Mar 20;23(6):3281. doi: 10.3390/s23063281. | 10.3390/s2306 3281 | 36991992 |
| Feasibility of smart wristbands for continuous monitoring during pregnancy and one month after birth | Grym K, Niela-Vilén H, Ekholm E, Hamari L, Azimi I, Rahmani A, Liljeberg P, Löyttyniemi E, Axelin A. | BMC Pregnancy Childbirth. 2019 Jan 17;19(1):34. doi: 10.1186/s12884-019-2187-9. | 10.1186/s12884-019-2187-9 | 30654747 |
| Quantifying physical activity in early Parkinson disease using a commercial activity monitor | Pradhan S, Kelly VE. | Parkinsonism Relat Disord. 2019 Sep;66:171-175. doi: 10.1016/j.parkreldis.2019.08.001. Epub 2019 Aug 3. | 10.1016/j.parkre ldis.2019.08.00 1 | 31420310 |
| Realization and Technology Acceptance Test of a Wearable Cardiac Health Monitoring and Early Warning System with Multi-Channel MCGs and ECG | Lin WY, Ke HL, Chou WC, Chang PC, Tsai TH, Lee MY. | Sensors (Basel). 2018 Oct 19;18(10):3538. doi: 10.3390/s18103538. | 10.3390/s18103 538 | 30347695 |
| Effect of Fitbit and iPad Wearable Technology in Health-Related Quality of | Yurkiewicz IR, Simon P, Liedtke M, Dahl G, Dunn T. | J Adolesc Young Adult Oncol. 2018 Oct;7(5):579-583. doi: | 10.1089/jayao.2 018.0022 | 29924668 |

| | | | | |
|---|---|---|---|---|
| Life in Adolescent and Young Adult Cancer Patients | | 10.1089/jayao.2018.0022. Epub 2018 Jun 20. | | |
| Parental Perspectives of a Wearable Activity Tracker for Children Younger Than 13 Years: Acceptability and Usability Study | Mackintosh KA, Chappel SE, Salmon J, Timperio A, Ball K, Brown H, Macfarlane S, Ridgers ND. | JMIR Mhealth Uhealth. 2019 Nov 4;7(11):e13858. doi: 10.2196/13858. | 10.2196/13858 | 31682585 |
| Feasibility of a Sensor-Based Technological Platform in Assessing Gait and Sleep of In-Hospital Stroke and Incomplete Spinal Cord Injury (iSCI) Patients | Hendriks MMS, Vos-van der Hulst M, Keijsers NLW. | Sensors (Basel). 2020 May 12;20(10):2748. doi: 10.3390/s20102748. | 10.3390/s20102748 | 32408490 |
| Community-Based ECG Monitoring System for Patients with Cardiovascular Diseases | Lin BS, Wong AM, Tseng KC. | J Med Syst. 2016 Apr;40(4):80. doi: 10.1007/s10916-016-0442-4. Epub 2016 Jan 22. | 10.1007/s10916-016-0442-4 | 26802010 |
| Home rehabilitation supported by a wearable soft-robotic device for improving hand function in older adults: A pilot randomized controlled trial | Radder B, Prange-Lasonder GB, Kottink AIR, Holmberg J, Sletta K, van Dijk M, Meyer T, Melendez-Calderon A, Buurke JH, Rietman JS. | PLoS One. 2019 Aug 6;14(8):e0220544. doi: 10.1371/journal.pone.0220544. eCollection 2019. | 10.1371/journal.pone.0220544 | 31386685 |
| Verification of a Portable Motion Tracking System for Remote Management of Physical Rehabilitation of the Knee | Bell KM, Onyeukwu C, McClincy MP, Allen M, Bechard L, Mukherjee A, Hartman RA, Smith C, Lynch AD, Irrgang JJ. | Sensors (Basel). 2019 Feb 28;19(5):1021. doi: 10.3390/s19051021. | 10.3390/s19051021 | 30823373 |
| Novel Bluetooth-Enabled Tubeless Insulin Pump: A User Experience Design Approach for a Connected Digital Diabetes Management Platform | Pillalamarri SS, Huyett LM, Abdel-Malek A. | J Diabetes Sci Technol. 2018 Nov;12(6):1132-1142. doi: 10.1177/1932296818804802. Epub 2018 Oct 11. | 10.1177/1932296818804802 | 30304951 |
| The mobile sleep lab app: An open-source framework for mobile sleep assessment based on consumer-grade wearable devices | Burgdorf A, Güthe I, Jovanović M, Kutafina E, Kohlschein C, Bitsch JÁ, Jonas SM. | Comput Biol Med. 2018 Dec 1;103:8-16. doi: 10.1016/j.compbiomed.2018.09.025. Epub 2018 Oct 6. | 10.1016/j.compbiomed.2018.09.025 | 30316065 |
| Comparison of Collaborative and Cooperative Schemes in Sensor Networks for Non-Invasive Monitoring of People at Home | Del-Valle-Soto C, Valdivia LJ, López-Pimentel JC, Visconti P. | Int J Environ Res Public Health. 2023 Mar 27;20(7):5268. doi: 10.3390/ijerph20075268. | 10.3390/ijerph20075268 | 37047884 |
| Feasibility of a Secure Wireless Sensing Smartwatch Application for the Self-Management of Pediatric Asthma | Hosseini A, Buonocore CM, Hashemzadeh S, Hojaiji H, Kalantarian H, Sideris C, Bui AAT, King CE, Sarrafzadeh M. | Sensors (Basel). 2017 Aug 3;17(8):1780. doi: 10.3390/s17081780. | 10.3390/s17081780 | 28771168 |
| Development of a Wearable Instrumented Vest for Posture Monitoring and System Usability Verification Based on the Technology Acceptance Model | Lin WY, Chou WC, Tsai TH, Lin CC, Lee MY. | Sensors (Basel). 2016 Dec 17;16(12):2172. doi: 10.3390/s16122172. | 10.3390/s16122172 | 27999324 |
| Preferred Placement and Usability of a Smart Textile System vs. Inertial Measurement Units for Activity Monitoring | Mokhlespour Esfahani MI, Nussbaum MA. | Sensors (Basel). 2018 Aug 1;18(8):2501. doi: 10.3390/s18082501. | 10.3390/s18082501 | 30071635 |

| | | | | |
|---|---|---|---|---|
| Biofeedback Treatment App for Pediatric Migraine: Development and Usability Study | Stubberud A, Tronvik E, Olsen A, Gravdahl G, Linde M. | Headache. 2020 May;60(5):889-901. doi: 10.1111/head.13772. Epub 2020 Feb 13. | 10.1111/head.13772 | 32052871 |
| Community-dwelling older adults' acceptance of smartwatches for health and location tracking | Chung J, Brakey HR, Reeder B, Myers O, Demiris G. | Int J Older People Nurs. 2023 Jan;18(1):e12490. doi: 10.1111/opn.12490. Epub 2022 Jul 12. | 10.1111/opn.12490 | 35818900 |
| A formative study exploring perceptions of physical activity and physical activity monitoring among children and young people with cystic fibrosis and health care professionals | Shelley J, Fairclough SJ, Knowles ZR, Southern KW, McCormack P, Dawson EA, Graves LEF, Hanlon C. | BMC Pediatr. 2018 Oct 23;18(1):335. doi: 10.1186/s12887-018-1301-x. | 10.1186/s12887-018-1301-x | 30352564 |
| Application of Activity Trackers among Nursing Home Residents-A Pilot and Feasibility Study on Physical Activity Behavior, Usage Behavior, Acceptance, Usability and Motivational Impact | Auerswald T, Meyer J, von Holdt K, Voelcker-Rehage C. | Int J Environ Res Public Health. 2020 Sep 14;17(18):6683. doi: 10.3390/ijerph17186683. | 10.3390/ijerph17186683 | 32937840 |
| Multimodal Remote Home Monitoring of Lung Transplant Recipients during COVID-19 Vaccinations: Usability Pilot Study of the COVIDA Desk Incorporating Wearable Devices | Schuurmans MM, Muszynski M, Li X, Marcinkevičs R, Zimmerli L, Monserrat Lopez D, Michel B, Weiss J, Hage R, Roeder M, Vogt JE, Brunschwiler T. | Medicina (Kaunas). 2023 Mar 20;59(3):617. doi: 10.3390/medicina59030617. | 10.3390/medicina59030617 | 36984618 |
| A Wearable Device for Breathing Frequency Monitoring: A Pilot Study on Patients with Muscular Dystrophy | Cesareo A, Nido SA, Biffi E, Gandossini S, D'Angelo MG, Aliverti A. | Sensors (Basel). 2020 Sep 18;20(18):5346. doi: 10.3390/s20185346. | 10.3390/s20185346 | 32961986 |
| An Integrated Multi-Sensor Approach for the Remote Monitoring of Parkinson's Disease | Albani G, Ferraris C, Nerino R, Chimienti A, Pettiti G, Parisi F, Ferrari G, Cau N, Cimolin V, Azzaro C, Priano L, Mauro A. | Sensors (Basel). 2019 Nov 2;19(21):4764. doi: 10.3390/s19214764. | 10.3390/s19214764 | 31684020 |
| Older Adults' Experience with a Novel Fall Detection Device | Demiris G, Chaudhuri S, Thompson HJ. | Telemed J E Health. 2016 Sep;22(9):726-32. doi: 10.1089/tmj.2015.0218. Epub 2016 Mar 9. | 10.1089/tmj.2015.0218 | 26959299 |
| Acceptability and utility of the mySentry remote glucose monitoring system | Kaiserman K, Buckingham BA, Prakasam G, Gunville F, Slover RH, Wang Y, Nguyen X, Welsh JB. | J Diabetes Sci Technol. 2013 Mar 1;7(2):356-61. doi: 10.1177/193229681300700211. | 10.1177/193229681300700211 | 23566993 |
| Using the Technology Acceptance Model to Explore Adolescents' Perspectives on Combining Technologies for Physical Activity Promotion Within an Intervention: Usability Study | Drehlich M, Naraine M, Rowe K, Lai SK, Salmon J, Brown H, Koorts H, Macfarlane S, Ridgers ND. | J Med Internet Res. 2020 Mar 6;22(3):e15552. doi: 10.2196/15552. | 10.2196/15552 | 32141834 |
| A qualitative evaluation of breast cancer survivors' acceptance of and preferences for consumer wearable technology activity trackers | Nguyen NH, Hadgraft NT, Moore MM, Rosenberg DE, Lynch C, Reeves MM, Lynch BM. | Support Care Cancer. 2017 Nov;25(11):3375-3384. doi: 10.1007/s00520-017-3756-y. Epub 2017 May 24. | 10.1007/s00520-017-3756-y | 28540402 |
| Multimodal nocturnal seizure detection in | Arends J, Thijs RD, Gutter T, Ungureanu C, | Neurology. 2018 Nov | 10.1212/WNL.00 | 30355702 |

| | | | | |
|---|---|---|---|---|
| a residential care setting: A long-term prospective trial | Cluitmans P, Van Dijk J, van Andel J, Tan F, de Weerd A, Vledder B, Hofstra W, Lazeron R, van Thiel G, Roes KCB, Leijten F; the Dutch Tele-Epilepsy Consortium. | 20;91(21):e2010-e2019. doi: 10.1212/WNL.0000000000006545. Epub 2018 Oct 24. | 000000000065 45 | |
| Feasibility of Interactive Resistance Chair in Older Adults with Diabetes | Finkelstein J, Cisse P, Jeong IC. | Stud Health Technol Inform. 2015;213:61-4. | | 26152953 |
| Usability of a wearable fall detection prototype from the perspective of older people-A real field testing approach | Thilo FJS, Hahn S, Halfens RJG, Schols JMGA. | J Clin Nurs. 2019 Jan;28(1-2):310-320. doi: 10.1111/jocn.14599. Epub 2018 Aug 1. | 10.1111/jocn.145 99 | 29964344 |
| Older Adults' Acceptance of Activity Trackers | Preusse KC, Mitzner TL, Fausset CB, Rogers WA. | J Appl Gerontol. 2017 Feb;36(2):127-155. doi: 10.1177/0733464815624151. Epub 2016 Jul 7. | 10.1177/073346 4815624151 | 26753803 |
| How is the usability of commercial activity monitors perceived by older adults and by researchers? A cross-sectional evaluation of community-living individuals | Hofbauer LM, Rodriguez FS. | BMJ Open. 2022 Nov 2;12(11):e063135. doi: 10.1136/bmjopen-2022-063135. | 10.1136/bmjope n-2022-063135 | 36323474 |
| StraightenUp+: Monitoring of Posture during Daily Activities for Older Persons Using Wearable Sensors | Cajamarca G, Rodríguez I, Herskovic V, Campos M, Riofrío JC. | Sensors (Basel). 2018 Oct 11;18(10):3409. doi: 10.3390/s18103409. | 10.3390/s18103 409 | 30314352 |
| Patient Engagement in Medical Device Design: Refining the Essential Attributes of a Wearable, Pre-Void, Ultrasound Alarm for Nocturnal Enuresis | Caswell N, Kuru K, Ansell D, Jones MJ, Watkinson BJ, Leather P, Lancaster A, Sugden P, Briggs E, Davies C, Oh C, Bennett K, DeGoede C. | Pharmaceut Med. 2020 Feb;34(1):39-48. doi: 10.1007/s40290-019-00324-w. | 10.1007/s40290 -019-00324-w | 31970684 |
| Multisensory Cues for Gait Rehabilitation with Smart Glasses: Methodology, Design, and Results of a Preliminary Pilot | Imbesi S, Corzani M. | Sensors (Basel). 2023 Jan 12;23(2):874. doi: 10.3390/s23020874. | 10.3390/s2302 0874 | 36679671 |
| Exoscore: A Design Tool to Evaluate Factors Associated With Technology Acceptance of Soft Lower Limb Exosuits by Older Adults | Shore L, Power V, Hartigan B, Schülein S, Graf E, de Eyto A, O'Sullivan L. | Hum Factors. 2020 May;62(3):391-410. doi: 10.1177/0018720819868122. Epub 2019 Aug 16. | 10.1177/0018720 819868122 | 31419179 |
| The design and evaluation of an activity monitoring user interface for people with stroke | Hart P, Bierwirth R, Fulk G, Sazonov E. | Annu Int Conf IEEE Eng Med Biol Soc. 2014;2014:5908-11. doi: 10.1109/EMBC.2014.6944973. | 10.1109/EMBC.2 014.6944973 | 25571341 |
| The role of individual differences on perceptions of wearable fitness device trust, usability, and motivational impact | Rupp MA, Michaelis JR, McConnell DS, Smither JA. | Appl Ergon. 2018 Jul;70:77-87. doi: 10.1016/j.apergo.2018.02.005. Epub 2018 Feb 21. | 10.1016/j.apergo .2018.02.005 | 29866329 |
| Implementation of Medication Event Reminder Monitors among patients diagnosed with drug susceptible tuberculosis in rural Viet Nam: A qualitative study | Drabarek D, Anh NT, Nhung NV, Hoa NB, Fox GJ, Bernays S. | PLoS One. 2019 Jul 22;14(7):e0219891. doi: 10.1371/journal.pone.0219891. eCollection 2019. | 10.1371/journal. pone.0219891 | 31329610 |
| The Effectiveness of a Web-Based | Vandelanotte C, Duncan MJ, Maher CA, | J Med Internet Res. 2018 Dec | 10.2196/11321 | 30563808 |

| | | | | |
|---|---|---|---|---|
| Computer-Tailored Physical Activity Intervention Using Fitbit Activity Trackers: Randomized Trial | Schoeppe S, Rebar AL, Power DA, Short CE, Doran CM, Hayman MJ, Alley SJ. | 18;20(12):e11321. doi: 10.2196/11321. | | |
| On-site Evaluation of Rehabilitation Patients Monitoring System Using Distributed Wireless Gateways | Matsunaga K, Ogasawara T, Kodate J, Mukaino M, Saitoh E. | Annu Int Conf IEEE Eng Med Biol Soc. 2019 Jul;2019:3195-3198. doi: 10.1109/EMBC.2019.8856963. | 10.1109/EMBC.2019.8856963 | 31946567 |
| ChroniSense National Early Warning Score Study: Comparison Study of a Wearable Wrist Device to Measure Vital Signs in Patients Who Are Hospitalized | Van Velthoven MH, Oke J, Kardos A. | J Med Internet Res. 2023 Feb 6;25:e40226. doi: 10.2196/40226. | 10.2196/40226 | 36745491 |
| Enhancing Visual Exploration through Augmented Gaze: High Acceptance of Immersive Virtual Biking by Oldest Olds | de'Sperati C, Dalmasso V, Moretti M, Høeg ER, Baud-Bovy G, Cozzi R, Ippolito J. | Int J Environ Res Public Health. 2023 Jan 17;20(3):1671. doi: 10.3390/ijerph20031671. | 10.3390/ijerph20031671 | 36767037 |
| Feasibility of a wearable soft-robotic glove to support impaired hand function in stroke patients | Radder B, Prange-Lasonder G, Kottink AIR, Melendez-Calderon A, Buurke JH, Rietman JS. | J Rehabil Med. 2018 Jul 17;50(7):598-606. doi: 10.2340/16501977-2357. | 10.2340/16501977-2357 | 30003268 |
| Measurement of daily physical activity using the SenseWear Armband: Compliance, comfort, adverse side effects and usability | McNamara RJ, Tsai LL, Wootton SL, Ng LW, Dale MT, McKeough ZJ, Alison JA. | Chron Respir Dis. 2016 May;13(2):144-54. doi: 10.1177/1479972316631138. Epub 2016 Feb 15. | 10.1177/1479972316631138 | 26879695 |
| The effect of a wearable soft-robotic glove on motor function and functional performance of older adults | Radder B, Prange-Lasonder GB, Kottink AIR, Holmberg J, Sletta K, Van Dijk M, Meyer T, Buurke JH, Rietman JS. | Assist Technol. 2020;32(1):9-15. doi: 10.1080/10400435.2018.1453888. Epub 2018 May 24. | 10.1080/10400435.2018.1453888 | 29601251 |

### Supplementary Table 3: Individual conditions falling into each therapeutic area

| **Therapeutic area** | **Disease or condition of study** |
|---|---|
| Aging | Older adults; aging adults; elderly. |
| Cardiovascular | Stroke including transient ischemic attack and minor ischemic stroke; heart failure; heart disease; hypertension. |
| Endocrine | Diabetes type 1; diabetes type 2; diabetes not further specified. |
| Neurology | Epilepsy; headaches; Parkinson's disease; major depressive disorder; glaucoma; seizures; traumatic brain injury. |
| Oncology | Any form of cancer. |
| Respiratory | Chronic obstructive pulmonary disease; cystic fibrosis; dust-related respiratory condition; lung transplant recipients; asthma; tuberculosis. |
| Surgery | Elective abdominal surgery; knee rehabilitation following injury or surgery; knee replacement surgery; post-operative patients. |
| Healthy | No specific disease or condition |
| Other | Anaphylaxis, muscular dystrophy, hemophilia, nocturnal enuresis, blood and marrow transplant, overweight/obesity, pregnancy, and non-specific hospitalized or chronic illness.One study recruited clinicians only (no end-users of the sDHT) and is included in this category. |

### Supplementary Table 4: Study design and sample characteristics across therapeutic areas; complete wear locations and health concepts

*This table aligns to Table 4 of the manuscript, and presents both the complete and collapsed lists of wear locations and health concepts across all studies.*

| | ***Therapeutic area of sDHT users*** | | | | | | | | | |
|---|---|---|---|---|---|---|---|---|---|---|
| | **Aging**<br>**27 sDHTs** | **Cardiovascular**<br>**12 sDHTs** | **Endocrine**<br>**3 sDHTs** | **Neurology**<br>**31 sDHTs** | **Oncology**<br>**8 sDHTs** | **Respiratory**<br>**15 sDHTs** | **Surgery**<br>**7 sDHTs** | **Healthy**<br>**35 sDHTs** | **Other [a]**<br>**26 sDHTs** | **Total**<br>**164 sDHTs** |
| **sDHT type** | | | | | | | | | | |
| *Wearable; n* | 26 | 9 | 1 | 30 | 8 | 11 | 7 | 33 | 16 | 141 |
| *Ambient; n* | 1 | 3 | 2 | 1 | | 4 | | 2 | 10 | 23 |
| **sDHT maturity** | | | | | | | | | | |
| *Prototype; n* | 9 | 5 | 1 | 5 | | | 2 | 5 | 11 | 38 |
| *Final or marketed; n* | 18 | 6 | 2 | 26 | 8 | 15 | 5 | 27 | 12 | 119 |
| *Not reported; n* | | 1 | | | | | | 3 | 3 | 7 |
| **Form factor** | | | | | | | | | | |
| *Adhesive patch; n* | 2 | | 1 | 5 | | | 1 | 2 | 1 | 12 |
| *Balance board; n* | | | | | | | | | 1 | 1 |
| *Camera; video or still; n* | | 1 | | | | | | | 2 | 3 |
| *Clip; n* | 4 | | | | | 1 | 1 | 1 | | 7 |
| *Clothing or shoes; n* | 5 | 3 | | | | | | 4 | 5 | 17 |
| *Contact lens; n* | | | | 1 | | | | | | 1 |
| *Cuff/wrap; n* | | 1 | | 1 | | | 1 | 2 | | 5 |
| *Electrode/s; n* | | | | 1 | | | | 2 | | 3 |
| *Exercise equipment; n* | | | 1 | | | | | | 2 | 3 |
| *Glasses; n* | 1 | | | | | | | 1 | | 2 |
| *Gloves; n* | 2 | 1 | | 1 | | | | | | 4 |
| *Glucometer; n* | | | 1 | | | | | | | 1 |

| | | | | | | | | | | |
|---|---|---|---|---|---|---|---|---|---|---|
| *Handheld thermometer; n* | | | | | | 1 | | | | 1 |
| *Mattress pad; n* | | 1 | | 1 | | | | | | 2 |
| *Medication package; n* | | | | | | 1 | | | 1 | 2 |
| *Phone or tablet; n* | | | | | | | | 1 | 4 | 5 |
| *Probe; n* | | | | 1 | | | | | 1 | 2 |
| *Ring; n* | | | | | | 1 | | | | 1 |
| *Spirometer; n* | | | | | | 2 | | | | 2 |
| *Contactless unit; n* | 1 | | | | | | | 1 | | 2 |
| *Strap; n* | 12 | 4 | | 20 | 8 | 9 | 4 | 21 | 9 | 87 |
| *Weight scale; n* | | 1 | | | | | | | | 1 |
| **Wear location** | | | | | | | | | | |
| *Arms/wrists/hands; n* | 11 | 5 | 0 | 19 | 8 | 8 | 3 | 19 | 9 | 82 |
| *Head/face; n* | 1 | 0 | 0 | 4 | 0 | 0 | 0 | 3 | 0 | 8 |
| *Legs/ankles/feet; n* | 2 | 2 | 0 | 1 | 0 | 0 | 2 | 3 | 0 | 10 |
| *Neck/torso/hips; n* | 10 | 2 | 1 | 3 | 0 | 2 | 2 | 6 | 6 | 32 |
| *Multiple locations; n* [b] | 2 | | | 3 | | | | 2 | 1 | 8 |
| *Not applicable; n* [c] | 1 | 3 | 2 | 1 | | 5 | | 2 | 10 | 24 |
| **Wear location** | | | | | | | | | | |
| *Ankle/s; n* | | 1 | | 1 | | | | | | 2 |
| *Arm; n* | 1 | 1 | | 6 | 1 | | 1 | 2 | | 12 |
| *Chest/torso/trunk; n* | 6 | 2 | 1 | 2 | | 1 | 1 | 4 | 6 | 23 |
| *Ear/s; n* | | | | | | | | 1 | | 1 |
| *Eye/s; n* | 1 | | | 1 | | | | 1 | | 3 |
| *Finger/s; n* | | | | 1 | | | 1 | | | 2 |
| *Foot/feet; n* | 1 | 1 | | | | | | 2 | | 4 |
| *Hand/s; n* | 1 | 1 | | 1 | | 1 | | | | 4 |
| *Head/scalp; n* | | | | 3 | | | | 1 | | 4 |
| *Hip/s; n* | 3 | | | | | 1 | | 1 | | 5 |
| *Leg/s; n* | 1 | | | | | | 2 | 1 | | 4 |

| | | | | | | | | | | |
|---|---|---|---|---|---|---|---|---|---|---|
| *Neck; n* | | | | | | | 1 | | | 1 |
| *Waist; n* | 1 | | | 1 | | | | 1 | | 3 |
| *Wrist; n* | 9 | 3 | | 11 | 7 | 7 | 1 | 17 | 9 | 64 |
| *Multiple locations; n* [b] | 2 | | | 3 | | | | 2 | 1 | 8 |
| *Not applicable; n* [c] | 1 | 3 | 2 | 1 | | 5 | | 2 | 10 | 24 |
| **Interaction type** [d] | | | | | | | | | | |
| *Passive; n* | 24 | 7 | 1 | 26 | 8 | 12 | 3 | 30 | 15 | 126 |
| *Active; n* | 3 | 5 | 2 | 5 | | 3 | 4 | 5 | 11 | 38 |
| **Health concepts** [e] | | | | | | | | | | |
| *Activities of daily living; n* | 6 | 2 | | 2 | | | | 1 | | 11 |
| *Physical activity; n* | 16 | 4 | 1 | 6 | 9 | 9 | 1 | 15 | 0 | 61 |
| *Adherence; n* | | | | | | 1 | | | | 1 |
| *Electrical activity; n* | 0 | 0 | 0 | 15 | 0 | 0 | 0 | 1 | 0 | 16 |
| *Mobility; n* | 5 | 4 | 0 | 11 | 0 | 0 | 2 | 13 | 0 | 35 |
| *Sleep; n* | 6 | 1 | 0 | 0 | 1 | 1 | 0 | 5 | 0 | 14 |
| *Vital signs; n* | 9 | 5 | 2 | 18 | 0 | 5 | 14 | 23 | 0 | 76 |
| *Other; n* [f] | 4 | 2 | 0 | 4 | 0 | 2 | 0 | 0 | 0 | 12 |
| **Health concepts** [e] | | | | | | | | | | |
| *Activities of daily living; n* | 6 | 2 | | 2 | | | | 1 | | 11 |
| *Adherence to therapy; n* | | | | | | 1 | | | | 1 |
| *Bladder volume; n* | | | | | | | | | | 0 |
| *Blood glucose; n* | | | 2 | | | | | | | 2 |
| *Blood oxygen; n* | 1 | | | | | 1 | 3 | 6 | | 11 |
| *Blood pressure, volume and/or arterial stiffness; n* | | 2 | | 5 | | | 3 | 1 | | 11 |
| *Body habitus; n* | | 1 | | | | | | | | 1 |
| *Cardiac output; n* | | 1 | | | | | | | | 1 |
| *Electrical activity or abnormalities including seizures; n* | | | | 11 | | | | 1 | | 12 |
| *Electrodermal activity; n* | | | | 4 | | | | | | 4 |

| | | | | | | | | | | |
|---|---|---|---|---|---|---|---|---|---|---|
| *Energy expenditure; n* | 3 | 1 | | | 1 | 1 | | 1 | | 7 |
| *Exercise; n* | 13 | 3 | 1 | 6 | 8 | 8 | 1 | 14 | | 54 |
| *Fall detection; n* | 3 | | | 1 | | | | | | 4 |
| *Foot pressure; n* | | | | | | | | 1 | | 1 |
| *Gait; n* | 3 | 1 | | 3 | | | | 2 | | 9 |
| *Gaze or visual movement; n* | 1 | | | | | | | | | 1 |
| *Heart rate/rhythm; n* | 6 | 2 | | 10 | | 2 | 4 | 11 | | 35 |
| *Intraocular pressure; n* | | | | 1 | | | | | | 1 |
| *Joint or head kinematics; n* | | 2 | | | | | 2 | 6 | | 10 |
| *Lung/airway function; n* | | | | | | 2 | | | | 2 |
| *Mobility; n* | 1 | 1 | | 8 | | | | 2 | | 12 |
| *Posture; n* | 1 | | | | | | | 2 | | 3 |
| *Respiratory rate, pattern, or drive; n* | 1 | | | | | | 2 | 5 | | 8 |
| *Sleep duration and/or continuity; n* | 6 | | | | 1 | 1 | | 3 | | 11 |
| *Sleep position or movement; n* | | | | | | | | 1 | | 1 |
| *Temperature; n* | 1 | 1 | | 3 | | 2 | 2 | | | 9 |
| *Tremor detection; n* | | | | 2 | | | | | | 2 |
| *Sleep staging; n* | | 1 | | | | | | 1 | | 2 |

[a] 'Other' therapeutic area category contains studies with enrollment eligibility focused on anaphylaxis, muscular dystrophy, hemophilia, nocturnal enuresis, blood and marrow transplant, overweight/obesity, pregnancy, and non-specific hospitalized or chronic illness.One study recruited clinicians only (no end-users of the sDHT) and is included in this category.

[b] Refers to multi-sensor sDHTs worn on different parts of the body, or sDHTs that can be positioned in one of many locations

[c] Wear location is not applicable to ambient sDHTs.

[d] Passive: sDHT data are collected over long time periods without user input other than aspects such as charging or changing batteries (such as actigraphy); includes tools for which the absence of data is meaningful (such as smart packaging for adherence monitoring). Active: sDHT data collection requires user engagement at defined timepoints.

[e] Health concepts are not mutually exclusive; a single sDHT can capture data in multiple categories.

[f] ‘Other’ health concept categories are described below.

### Supplementary Table 5: Study design and sample characteristics across therapeutic areas; Summative studies

*This table aligns to Table 3 of the manuscript, and presents data from studies reporting summative evaluations only and studies reporting both summative and formative evaluations.*

| | ***Therapeutic area of sDHT end-users*** | | | | | | | | | |
|---|---|---|---|---|---|---|---|---|---|---|
| | ***Aging*** **9 studies** | ***Cardiovascular*** **5 studies** | ***Endocrine*** **2 studies** | ***Neurology*** **11 studies** | ***Oncology*** **3 studies** | ***Respiratory*** **5 studies** | ***Surgery*** **3 studies** | ***Healthy*** **10 studies** | ***Other*** [a] **7 studies** | ***Total*** **55 studies** |
| ***Study Design*** | | | | | | | | | | |
| *Observational; n* | 9 | 5 | 2 | 11 | 3 | 4 | 2 | 9 | 7 | 52 |
| *Interventional; n* | | | | | | 1 | 1 | 1 | | 3 |
| ***Study Focus*** [b] | | | | | | | | | | |
| *Summative; sample size rationale n* | 3 | 3 | | 4 | | 1 | 2 | 1 | 3 | 17 |
| *Summative; no sample size rationale n* | 6 | 2 | 2 | 7 | 3 | 4 | 1 | 9 | 4 | 38 |
| *Formative; sample size rationale n* | | | | | | | | | | 0 |
| *Formative; no sample size rationale n* | | | | | | | | | | 0 |
| ***Setting*** | | | | | | | | | | |
| *Remote; n* | 5 | 3 | 1 | 5 | 2 | 5 | | 6 | 3 | 30 |
| *On-site; n* | 2 | 2 | | 4 | | | 2 | 4 | 3 | 17 |
| *Both remote and on-site; n* | 2 | | 1 | 2 | 1 | | 1 | | 1 | 8 |
| ***Duration of sDHT data collection*** | | | | | | | | | | |
| *≤1 day; n* | 1 | 1 | | 1 | | | 1 | 3 | | 7 |
| *>1, ≤7 days; n* | 1 | 1 | | 4 | | 2 | 2 | 2 | 2 | 14 |
| *>7, ≤30 days; n* | 3 | 2 | 1 | 2 | 1 | | | 2 | | 11 |
| *>31, ≤90 days; n* | 3 | | | 2 | 1 | 3 | | 2 | | 11 |
| *>90, ≤180 days; n* | | 1 | | | 1 | | | | 3 | 5 |
| *>180 days; n* | 1 | | | 2 | | | | | 1 | 4 |
| *Not reported; n* | | | 1 | | | | | 1 | 1 | 3 |
| ***Study Sample*** | | | | | | | | | | |
| *Sample size;* | 30 (8 - 110) | 30 (5 - 156) | 189 (35 - | 60 (20 - | 30 (14 - 33) | 19 (9 - 314) | 60 (29 - | 77 (5 - 243) | 20 (10 - | 30 (5 - |

| | | | | | | | | | | |
|---|---|---|---|---|---|---|---|---|---|---|
| *median (min - max)* | | | 343) | 623) | | | 77) | | 407) | 623) |
| *End-users; n* [c] | 9 | 5 | 2 | 11 | 3 | 5 | 3 | 10 | 6 | 54 |
| *Carepartner-users; n* [c] | | | 1 | 2 | | | | 3 | | 6 |
| *Clinician-users; n* [c] | 1 | 2 | 1 | 2 | | 1 | 1 | 1 | 1 | 10 |
| *Experts; n* [c] | | | 1 | | | | | 1 | | 2 |
| *Adults only; n* | 9 | 5 | | 9 | 2 | 3 | 3 | 6 | 5 | 42 |
| *Children only; n* | | | 1 | | | 1 | | 3 | 2 | 7 |
| *Both adults and children; n* | | | | 2 | 1 | 1 | | 1 | | 5 |
| *Not reported; n* | | | 1 | | | | | | | 1 |
| *Males/men only; n* | | | | | | | | | 2 | 2 |
| *Females/women only; n* | 1 | | | | 1 | | | | 1 | 3 |
| *Both or all sexes/genders; n* | 8 | 5 | 1 | 11 | 2 | 5 | 3 | 9 | 4 | 48 |
| *Not reported; n* | | | 1 | | | | | 1 | | 2 |
| *Race/ethnicity reported; n* | 1 | 1 | | 4 | | 1 | | 2 | 1 | 10 |
| *Race/ethnicity not reported; n* | 8 | 4 | 2 | 7 | 3 | 4 | 3 | 8 | 6 | 45 |
| ***Number of sDHTs assessed*** | | | | | | | | | | |
| *Range (min - max)* | 1 - 7 | 1 - 3 | 1 - 1 | 1 - 5 | 1 - 6 | 1 - 5 | 1 - 2 | 1 - 7 | 1 - 11 | 1 - 11 |

[a] 'Other' therapeutic area category contains studies with enrollment eligibility focused on anaphylaxis, muscular dystrophy, hemophilia, nocturnal enuresis, blood and marrow transplant, overweight/obesity, pregnancy, and non-specific hospitalized or chronic illness. One study recruited clinicians only (no end-users of the sDHT) and is included in this category.

[b] Studies reporting formative evaluations only are described in Supplementary Table 8.

[c] Categories are not mutually-exclusive

### Supplementary Table 6: Sensor-based digital health technology descriptive information across therapeutic areas; Summative studies

*This table aligns to Table 4 of the manuscript, and presents data from studies reporting summative evaluations only and studies reporting both summative and formative evaluations. Complete lists of wear locations and health concepts are provided, aligned to Supplementary Table 4.*

| | ***Therapeutic area of sDHT users*** | | | | | | | | | |
|---|---|---|---|---|---|---|---|---|---|---|
| | **Aging** | **Cardiovascular** | **Endocrine** | **Neurology** | **Oncology** | **Respiratory** | **Surgery** | **Healthy** | **Other [a]** | **Total** |
| | ***9 studies*** | ***5 studies*** | ***2 studies*** | ***11 studies*** | ***3 studies*** | ***5 studies*** | ***3 studies*** | ***10 studies*** | ***7 studies*** | ***55 studies*** |
| **sDHT type** | | | | | | | | | | |
| *Wearable; n* | 17 | 6 | 1 | 25 | 8 | 10 | 5 | 26 | 14 | 112 |
| *Ambient; n* | | 1 | 1 | 1 | | 3 | | | 9 | 15 |
| **sDHT maturity** | | | | | | | | | | |
| *Prototype; n* | 1 | 2 | | 3 | | | | 1 | 8 | 15 |
| *Final or marketed; n* | 16 | 4 | 2 | 23 | 8 | 13 | 5 | 24 | 12 | 107 |
| *Not reported; n* | | 1 | | | | | | 1 | 3 | 5 |
| **Form factor** | | | | | | | | | | |
| *Adhesive patch; n* | 1 | | 1 | 4 | | | 1 | 2 | 1 | 10 |
| *Balance board; n* | | | | | | | | | 1 | 1 |
| *Camera; video or still; n* | | | | | | | | | 2 | 2 |
| *Clip; n* | 2 | | | | | 1 | 1 | 1 | | 5 |
| *Clothing or shoes; n* | 2 | 1 | | | | | | 3 | 4 | 10 |
| *Contact lens; n* | | | | 1 | | | | | | 1 |
| *Cuff/wrap; n* | | 1 | | 1 | | | 1 | 1 | | 4 |
| *Electrode/s; n* | | | | 1 | | | | 1 | | 2 |
| *Exercise equipment; n* | | | | | | | | | 2 | 2 |
| *Glasses; n* | | | | | | | | 1 | | 1 |
| *Gloves; n* | 1 | 1 | | | | | | | | 2 |
| *Glucometer; n* | | | 1 | | | | | | | 1 |

| | | | | | | | | | | |
|---|---|---|---|---|---|---|---|---|---|---|
| *Handheld thermometer; n* | | | | | | 1 | | | | 1 |
| *Mattress pad; n* | | | | 1 | | | | | | 1 |
| *Medication package; n* | | | | | | 1 | | | | 1 |
| *Phone or tablet; n* | | | | | | | | | 4 | 4 |
| *Probe; n* | | | | | | | | | | 0 |
| *Ring; n* | | | | | | 1 | | | | 1 |
| *Spirometer; n* | | | | | | 1 | | | | 1 |
| *Contactless unit; n* | | | | | | | | | | 0 |
| *Strap; n* | 11 | 3 | | 18 | 8 | 8 | 2 | 17 | 9 | 76 |
| *Weight scale; n* | | 1 | | | | | | | | 1 |
| **Wear location** | | | | | | | | | | |
| *Arms/wrists/hands; n* | 11 | 5 | 0 | 16 | 8 | 8 | 3 | 17 | 9 | 77 |
| *Head/face; n* | 0 | 0 | 0 | 4 | 0 | 0 | 0 | 2 | 0 | 6 |
| *Legs/ankles/feet; n* | | | | | | | | | | 5 |
| *Neck/torso/hips; n* | 5 | 0 | 1 | 3 | 0 | 2 | 2 | 4 | 5 | 22 |
| *Multiple locations; n* [b] | | | | 1 | | | | 1 | | 2 |
| *Not applicable; n* [c] | | 1 | 1 | 1 | | 3 | | | 9 | 15 |
| **Wear location** | | | | | | | | | | |
| *Ankle/s; n* | | | | 1 | | | | | | 1 |
| *Arm; n* | 1 | 1 | | 6 | 1 | | 1 | 1 | | 11 |
| *Chest/torso/trunk; n* | 2 | | 1 | 2 | | 1 | 1 | 3 | 5 | 15 |
| *Ear/s; n* | | | | | | | | 1 | | 1 |
| *Eye/s; n* | | | | 1 | | | | 1 | | 2 |
| *Finger/s; n* | | | | | | | 1 | | | 1 |
| *Foot/feet; n* | 1 | 1 | | | | | | 2 | | 4 |
| *Hand/s; n* | 1 | 1 | | | | 1 | | | | 3 |
| *Head/scalp; n* | | | | 3 | | | | | | 3 |
| *Hip/s; n* | 2 | | | | | 1 | | 1 | | 4 |
| *Leg/s; n* | | | | | | | | | | 0 |

| | | | | | | | | | | |
|---|---|---|---|---|---|---|---|---|---|---|
| *Neck; n* | | | | | | | 1 | | | 1 |
| *Waist; n* | 1 | | | 1 | | | | | | 2 |
| *Wrist; n* | 9 | 3 | | 10 | 7 | 7 | 1 | 16 | 9 | 62 |
| *Multiple locations; n* [b] | | | | 1 | | | | 1 | | 2 |
| *Not applicable; n* [c] | | 1 | 1 | 1 | | 3 | | | 9 | 15 |
| **Interaction type** [d] | | | | | | | | | | |
| *Passive; n* | 15 | 3 | 1 | 26 | 8 | 11 | 3 | 25 | 13 | 105 |
| *Active; n* | 2 | 4 | 1 | 0 | 0 | 2 | 2 | 1 | 10 | 22 |
| **Health concepts** [e] | | | | | | | | | | |
| *Activities of daily living; n* | 4 | 2 | | | | | | 1 | | 7 |
| *Physical activity; n* | 15 | 4 | 0 | 6 | 9 | 8 | 0 | 14 | 0 | 56 |
| *Adherence; n* | | | | | | 1 | | | | 1 |
| *Electrical activity; n* | 0 | 0 | 0 | 14 | 0 | 0 | 0 | 0 | 0 | 14 |
| *Mobility; n* | 1 | 2 | 0 | 9 | 0 | 0 | 0 | 7 | 0 | 19 |
| *Sleep; n* | 6 | 0 | 0 | 0 | 1 | 1 | 0 | 2 | 0 | 10 |
| *Vital signs; n* | 8 | 4 | 2 | 16 | 0 | 4 | 14 | 21 | 0 | 69 |
| *Other; n* [f] | 0 | 1 | 0 | 4 | 0 | 1 | 0 | 0 | 0 | 6 |
| **Health concepts** [e] | | | | | | | | | | |
| *Activities of daily living; n* | 4 | 2 | | | | | | 1 | | 7 |
| *Adherence to therapy; n* | | | | | | 1 | | | | 1 |
| *Bladder volume; n* | | | | | | | | | | 0 |
| *Blood glucose; n* | | | 2 | | | | | | | 2 |
| *Blood oxygen; n* | 1 | | | | | 1 | 3 | 6 | | 11 |
| *Blood pressure, volume and/or arterial stiffness; n* | | 2 | | 5 | | | 3 | 1 | | 11 |
| *Body habitus; n* | | 1 | | | | | | | | 1 |
| *Cardiac output; n* | | | | | | | | | | 0 |
| *Electrical activity or abnormalities including seizures; n* | | | | 10 | | | | | | 10 |
| *Electrodermal activity; n* | | | | 4 | | | | | | 4 |

| | | | | | | | | | | |
|---|---|---|---|---|---|---|---|---|---|---|
| *Energy expenditure; n* | 3 | 1 | | | 1 | 1 | | 1 | | 7 |
| *Exercise; n* | 12 | 3 | | 6 | 8 | 7 | | 13 | | 49 |
| *Fall detection; n* | | | | 1 | | | | | | 1 |
| *Foot pressure; n* | | | | | | | | 1 | | 1 |
| *Gait; n* | 1 | | | 2 | | | | 1 | | 4 |
| *Gaze or visual movement; n* | | | | | | | | | | 0 |
| *Heart rate/rhythm; n* | 5 | 1 | | 9 | | 1 | 4 | 9 | | 29 |
| *Intraocular pressure; n* | | | | 1 | | | | | | 1 |
| *Joint or head kinematics; n* | | 1 | | | | | | 2 | | 3 |
| *Lung/airway function; n* | | | | | | 1 | | | | 1 |
| *Mobility; n* | | 1 | | 7 | | | | 1 | | 9 |
| *Posture; n* | | | | | | | | 2 | | 2 |
| *Respiratory rate, pattern, or drive; n* | 1 | | | | | | 2 | 5 | | 8 |
| *Sleep duration and/or continuity; n* | 6 | | | | 1 | 1 | | 2 | | 10 |
| *Sleep position or movement; n* | | | | | | | | | | 0 |
| *Temperature; n* | 1 | 1 | | 2 | | 2 | 2 | | | 8 |
| *Tremor detection; n* | | | | 2 | | | | | | 2 |
| *Sleep staging; n* | | | | | | | | | | 0 |

[a] 'Other' therapeutic area category contains studies with enrollment eligibility focused on anaphylaxis, muscular dystrophy, hemophilia, nocturnal enuresis, blood and marrow transplant, overweight/obesity, pregnancy, and non-specific hospitalized or chronic illness.One study recruited clinicians only (no end-users of the sDHT) and is included in this category.

[b] Refers to multi-sensor sDHTs worn on different parts of the body, or sDHTs that can be positioned in one of many locations

[c] Wear location is not applicable to ambient sDHTs.

[d] Passive: sDHT data are collected over long time periods without user input other than aspects such as charging or changing batteries (such as actigraphy); includes tools for which the absence of data is meaningful (such as smart packaging for adherence monitoring). Active: sDHT data collection requires user engagement at defined timepoints.

[e] Health concepts are not mutually exclusive; a single sDHT can capture data in multiple categories.

[f] ‘Other’ health concept categories are described below.

### Supplementary Table 7: Methodological approaches to human factors data collection; Summative studies

*This table aligns to Table 5 of the manuscript, and presents data from studies reporting summative evaluations only and studies reporting both summative and formative evaluations.*

| | sDHT type | | |
|---|---|---|---|
| | **Ambient**<br>**15 sDHTs** | **Wearable**<br>**112 sDHTs** | **Total**<br>**127 sDHTs** |
| **Data collection environment** | | | |
| *Actual environment; n* | 15 | 88 | 103 |
| *Simulated environment; n* | | 19 | 19 |
| *Both actual and simulated; n* | | 5 | 5 |
| **Interactions with sDHT** | | | |
| *Look-and-feel; n* | | 12 | 12 |
| *Actual use; n* | 15 | 100 | 115 |
| **Human factors evaluation methods** [a] | | | |
| *Interviews; n* | 2 | 38 | 40 |
| *Focus groups; n* | 10 | 17 | 27 |
| *Surveys - referenced; n* | 11 | 55 | 66 |
| *Surveys - in-house; n* | 3 | 61 | 64 |
| *Think-aloud; n* | | 14 | 14 |
| *Observation (direct or video); n* | 1 | 23 | 24 |
| *Measured by the sDHT; n* | | 4 | 4 |
| *Heuristic analysis; n* | 1 | 1 | 2 |
| **Type/s of human factors data reported** | | | |
| *Mixed methods; n* | 12 | 53 | 65 |
| *Quantitative only; n* | 2 | 40 | 42 |
| *Qualitative only; n* | 1 | 19 | 20 |
| **Categories of human factors and related data reported** | | | 0 |

| | | | |
|---|---|---|---|
| *User satisfaction; n* | 14 | 94 | 108 |
| *Comfort; n* | 3 | 91 | 94 |
| *Ease of use; self-report; n* | 15 | 93 | 108 |
| *Ease of use; objectively-captured; n* | | 4 | 4 |
| *Learnability; n* | | 10 | 10 |
| *Efficiency; n* | | 4 | 4 |
| *Memorability; n* | | 2 | 2 |
| *Usefulness; n* | 11 | 77 | 88 |
| *Use errors; n* | 4 | 19 | 23 |
| *User trust; n* | 10 | 44 | 54 |
| *Information readability; n* | | | 0 |
| *Information understandability and/or actionability; n* | | 8 | 8 |
| *Technical performance or product-errors; n* | 13 | 68 | 81 |
| **Adherence to sDHT reported** | | | |
| *Objectively-measured by the sDHT; n* | 4 | 37 | 41 |
| *Self/carepartner report; n* | | 13 | 13 |
| *Both objective and self/carepartner; n* | | 4 | 4 |
| *Reported but method not described; n* | 1 | 6 | 7 |
| *Adherence not reported; n* | 10 | 52 | 62 |

[a] categories are not mutually exclusive

### Supplementary Table 8: Study design and sample characteristics across therapeutic areas; Formative studies

*This table aligns to Table 3 of the manuscript, and presents data from studies reporting formative evaluations only.*

| | **Therapeutic area of sDHT end-users** | | | | | | | | | |
|---|---|---|---|---|---|---|---|---|---|---|
| | ***Aging*** **10 studies** | ***Cardiovascular*** **4 studies** | ***Endocrine*** **1 studies** | ***Neurology*** **2 studies** | ***Oncology*** **0 studies** | ***Respiratory*** **1 studies** | ***Surgery*** **2 studies** | ***Healthy*** **5 studies** | ***Other*** [a] **3 studies** | ***Total*** **28 studies** |
| ***Study Design*** | | | | | | | | | | |
| *Observational; n* | 8 | 4 | 1 | 2 | | 1 | 2 | 5 | 3 | 26 |
| *Interventional; n* | 2 | | | | | | | | | 2 |
| ***Study Focus*** [b] | | | | | | | | | | |
| *Summative; sample size rationale n* | | | | | | | | | | 0 |
| *Summative; no sample size rationale n* | | | | | | | | | | 0 |
| *Formative; sample size rationale n* | 2 | 1 | | | | | | | 1 | 4 |
| *Formative; no sample size rationale n* | 8 | 3 | 1 | 2 | | 1 | 2 | 5 | 2 | 24 |
| ***Setting*** | | | | | | | | | | |
| *Remote; n* | 6 | | | | | 1 | 1 | 1 | 3 | 12 |
| *On-site; n* | 2 | 4 | 1 | 1 | | | 1 | 4 | | 13 |
| *Both remote and on-site; n* | 2 | | | 1 | | | | | | 3 |
| ***Duration of sDHT data collection*** | | | | | | | | | | |
| *≤1 day; n* | 4 | 2 | 1 | 1 | | | 1 | 4 | 2 | 15 |
| *>1, ≤7 days; n* | 1 | 2 | | | | | | | | 3 |
| *>7, ≤30 days; n* | 3 | | | 1 | | | 1 | | | 5 |
| *>31, ≤90 days; n* | | | | | | 1 | | 1 | 1 | 3 |
| *>90, ≤180 days; n* | 1 | | | | | | | | | 1 |
| *>180 days; n* | | | | | | | | | | 0 |
| *Not reported; n* | 1 | | | | | | | | | 1 |
| ***Study Sample*** | | | | | | | | | | |
| *Sample size; median (min - max)* | 23 (8 - 125) | 17 (5 - 48) | 5 (5 - 5) | 22.5 (5 - 40) | #N/A | 1 (1 - 1) | 12.5 (10 - 15) | 11 (1 - 50) | 22 (3 - 99) | 15 (1 - 125) |

| | | | | | | | | | | |
|---|---|---|---|---|---|---|---|---|---|---|
| *End-users; n* [c] | 10 | 4 | 1 | 2 | | 1 | 2 | 5 | 3 | 28 |
| *Carepartner-users; n* [c] | | 1 | | | | | | | 1 | 2 |
| *Clinician-users; n* [c] | 1 | 1 | | | | | | | | 2 |
| *Experts; n* [c] | 1 | | | | | | | | | 1 |
| *Adults only; n* | 10 | 2 | 1 | 1 | | | 2 | 5 | 2 | 23 |
| *Children only; n* | | | | 1 | | 1 | | | 1 | 3 |
| *Both adults and children; n* | | 1 | | | | | | | | 1 |
| *Not reported; n* | | 1 | | | | | | | | 1 |
| *Males/men only; n* | | | | | | | | 1 | | 1 |
| *Females/women only; n* | | | | | | 1 | | | 1 | 2 |
| *Both or all sexes/genders; n* | 10 | 2 | 1 | 2 | | | 2 | 4 | 2 | 23 |
| *Not reported; n* | | 2 | | | | | | | | 2 |
| *Race/ethnicity reported; n* | 1 | | | | | 1 | | 1 | 1 | 4 |
| *Race/ethnicity not reported; n* | 9 | 4 | 1 | 2 | | | 2 | 4 | 2 | 24 |
| ***Number of sDHTs assessed*** | | | | | | | | | | |
| *Range (min - max)* | 1 - 1 | 1 - 2 | 1 - 1 | 2 - 3 | #N/A | 2 - 2 | 1 - 1 | 1 - 5 | 1 - 1 | 1 - 5 |

[a] 'Other' therapeutic area category contains studies with enrollment eligibility focused on anaphylaxis, muscular dystrophy, hemophilia, nocturnal enuresis, blood and marrow transplant, overweight/obesity, pregnancy, and non-specific hospitalized or chronic illness. One study recruited clinicians only (no end-users of the sDHT) and is included in this category.

[b] Studies reporting summative evaluations only and both summative and formative evaluations are described in Supplementary Table 5.

[c] Categories are not mutually-exclusive

### Supplementary Table 9: Sensor-based digital health technology descriptive information across therapeutic areas; Formative studies

*This table aligns to Table 4 of the manuscript, and presents data from studies reporting formative evaluations only. Complete lists of wear locations and health concepts are provided, aligned to Supplementary Table 4.*

| | ***Therapeutic area of sDHT users*** | | | | | | | | | |
|---|---|---|---|---|---|---|---|---|---|---|
| | **Aging**<br>***10 studies*** | **Cardiovascular**<br>***4 studies*** | **Endocrine**<br>***1 studies*** | **Neurology**<br>***2 studies*** | **Oncology**<br>***0 studies*** | **Respiratory**<br>***1 studies*** | **Surgery**<br>***2 studies*** | **Healthy**<br>***5 studies*** | **Other [a]**<br>***3 studies*** | **Total**<br>***28 studies*** |
| **sDHT type** | | | | | | | | | | |
| *Wearable; n* | 9 | 3 | | 5 | | 1 | 2 | 7 | 2 | 29 |
| *Ambient; n* | 1 | 2 | 1 | | | 1 | | 2 | 1 | 8 |
| **sDHT maturity** | | | | | | | | | | |
| *Prototype; n* | 8 | 3 | 1 | 2 | | | 2 | 4 | 3 | 23 |
| *Final or marketed; n* | 2 | 2 | | 3 | | 2 | | 3 | | 12 |
| *Not reported; n* | | | | | | | | 2 | | 2 |
| **Form factor** | | | | | | | | | | |
| *Adhesive patch; n* | 1 | | | 1 | | | | | | 2 |
| *Balance board; n* | | | | | | | | | | 0 |
| *Camera; video or still; n* | | 1 | | | | | | | | 1 |
| *Clip; n* | 2 | | | | | | | | | 2 |
| *Clothing or shoes; n* | 3 | 2 | | | | | | 1 | 1 | 7 |
| *Contact lens; n* | | | | | | | | | | 0 |
| *Cuff/wrap; n* | | | | | | | | 1 | | 1 |
| *Electrode/s; n* | | | | | | | | 1 | | 1 |
| *Exercise equipment; n* | | | 1 | | | | | | | 1 |
| *Glasses; n* | 1 | | | | | | | | | 1 |
| *Gloves; n* | 1 | | | 1 | | | | | | 2 |
| *Glucometer; n* | | | | | | | | | | 0 |

| | | | | | | | | | | |
|---|---|---|---|---|---|---|---|---|---|---|
| *Handheld thermometer; n* | | | | | | | | | | 0 |
| *Mattress pad; n* | | 1 | | | | | | | | 1 |
| *Medication package; n* | | | | | | | | | 1 | 1 |
| *Phone or tablet; n* | | | | | | | | 1 | | 1 |
| *Probe; n* | | | | 1 | | | | | 1 | 2 |
| *Ring; n* | | | | | | | | | | 0 |
| *Spirometer; n* | | | | | | 1 | | | | 1 |
| *Contactless unit; n* | 1 | | | | | | | 1 | | 2 |
| *Strap; n* | 1 | 1 | | 2 | | 1 | 2 | 4 | | 11 |
| *Weight scale; n* | | | | | | | | | | 0 |
| **Wear location** | | | | | | | | | | |
| *Arms/wrists/hands; n* | 0 | 0 | 0 | 3 | | 0 | 0 | 2 | 0 | 5 |
| *Head/face; n* | | | | | | | | | | 2 |
| *Legs/ankles/feet; n* | | | | | | | | | | 5 |
| *Neck/torso/hips; n* | | | | | | | | | | 10 |
| *Multiple locations; n* [b] | 2 | | | 2 | | | | 1 | 1 | 6 |
| *Not applicable; n* [c] | 1 | 2 | 1 | | | 2 | | 2 | 1 | 9 |
| **Wear location** | | | | | | | | | | |
| *Ankle/s; n* | | 1 | | | | | | | | 1 |
| *Arm; n* | | | | | | | | 1 | | 1 |
| *Chest/torso/trunk; n* | 4 | 2 | | | | | | 1 | 1 | 8 |
| *Ear/s; n* | | | | | | | | | | 0 |
| *Eye/s; n* | 1 | | | | | | | | | 1 |
| *Finger/s; n* | | | | 1 | | | | | | 1 |
| *Foot/feet; n* | | | | | | | | | | 0 |
| *Hand/s; n* | | | | 1 | | | | | | 1 |
| *Head/scalp; n* | | | | | | | | 1 | | 1 |
| *Hip/s; n* | 1 | | | | | | | | | 1 |
| *Leg/s; n* | 1 | | | | | | 2 | 1 | | 4 |

| | | | | | | | | | | |
|---|---|---|---|---|---|---|---|---|---|---|
| *Neck; n* | | | | | | | | | | 0 |
| *Waist; n* | | | | | | | | 1 | | 1 |
| *Wrist; n* | | | | 1 | | | | 1 | | 2 |
| *Multiple locations; n* [b] | 2 | | | 2 | | | | 1 | 1 | 6 |
| *Not applicable; n* [c] | 1 | 2 | 1 | | | 2 | | 2 | 1 | 9 |
| **Interaction type** [d] | | | | | | | | | | |
| *Passive; n* | 9 | 4 | | | | 1 | | 5 | 2 | 21 |
| *Active; n* | 1 | 1 | 1 | 5 | | 1 | 2 | 4 | 1 | 16 |
| **Health concepts** [e] | | | | | | | | | | |
| *Activities of daily living; n* | 2 | | | 2 | | | | | | 4 |
| *Physical activity; n* | 1 | 0 | 1 | 0 | 0 | 1 | 1 | 1 | 0 | 5 |
| *Adherence; n* | | | | | | | | | | 0 |
| *Electrical activity; n* | 0 | 0 | 0 | 1 | 0 | 0 | 0 | 1 | 0 | 2 |
| *Mobility; n* | 4 | 2 | 0 | 2 | 0 | 0 | 2 | 6 | 0 | 16 |
| *Sleep; n* | 0 | 1 | 0 | 0 | 0 | 0 | 0 | 3 | 0 | 4 |
| *Vital signs; n* | 1 | 1 | 0 | 2 | 0 | 1 | 0 | 2 | 0 | 7 |
| *Other; n* [f] | 4 | 1 | 0 | 0 | 0 | 1 | 0 | 0 | 0 | 6 |
| **Health concepts** [e] | | | | | | | | | | |
| *Activities of daily living; n* | 2 | | | 2 | | | | | | 4 |
| *Adherence to therapy; n* | | | | | | | | | | 0 |
| *Bladder volume; n* | | | | | | | | | | 0 |
| *Blood glucose; n* | | | | | | | | | | 0 |
| *Blood oxygen; n* | | | | | | | | | | 0 |
| *Blood pressure, volume and/or arterial stiffness; n* | | | | | | | | | | 0 |
| *Body habitus; n* | | | | | | | | | | 0 |
| *Cardiac output; n* | | 1 | | | | | | | | 1 |
| *Electrical activity or abnormalities including seizures; n* | | | | 1 | | | | 1 | | 2 |
| *Electrodermal activity; n* | | | | | | | | | | 0 |

| | | | | | | | | | | |
|---|---|---|---|---|---|---|---|---|---|---|
| *Energy expenditure; n* | | | | | | | | | | 0 |
| *Exercise; n* | 1 | | 1 | | | 1 | 1 | 1 | | 5 |
| *Fall detection; n* | 3 | | | | | | | | | 3 |
| *Foot pressure; n* | | | | | | | | | | 0 |
| *Gait; n* | 2 | 1 | | 1 | | | | 1 | | 5 |
| *Gaze or visual movement; n* | 1 | | | | | | | | | 1 |
| *Heart rate/rhythm; n* | 1 | 1 | | 1 | | 1 | | 2 | | 6 |
| *Intraocular pressure; n* | | | | | | | | | | 0 |
| *Joint or head kinematics; n* | | 1 | | | | | 2 | 4 | | 7 |
| *Lung/airway function; n* | | | | | | 1 | | | | 1 |
| *Mobility; n* | 1 | | | 1 | | | | 1 | | 3 |
| *Posture; n* | 1 | | | | | | | | | 1 |
| *Respiratory rate, pattern, or drive; n* | | | | | | | | | | 0 |
| *Sleep duration and/or continuity; n* | | | | | | | | 1 | | 1 |
| *Sleep position or movement; n* | | | | | | | | 1 | | 1 |
| *Temperature; n* | | | | 1 | | | | | | 1 |
| *Tremor detection; n* | | | | | | | | | | 0 |
| *Sleep staging; n* | | 1 | | | | | | 1 | | 2 |

[a] 'Other' therapeutic area category contains studies with enrollment eligibility focused on anaphylaxis, muscular dystrophy, hemophilia, nocturnal enuresis, blood and marrow transplant, overweight/obesity, pregnancy, and non-specific hospitalized or chronic illness.One study recruited clinicians only (no end-users of the sDHT) and is included in this category.

[b] Refers to multi-sensor sDHTs worn on different parts of the body, or sDHTs that can be positioned in one of many locations

[c] Wear location is not applicable to ambient sDHTs.

[d] Passive: sDHT data are collected over long time periods without user input other than aspects such as charging or changing batteries (such as actigraphy); includes tools for which the absence of data is meaningful (such as smart packaging for adherence monitoring). Active: sDHT data collection requires user engagement at defined timepoints.

[e] Health concepts are not mutually exclusive; a single sDHT can capture data in multiple categories.

[f] ‘Other’ health concept categories are described below.

### Supplementary Table 10: Methodological approaches to human factors data collection; Formative studies

*This table aligns to Table 5 of the manuscript, and presents data from studies reporting formative evaluations only..*

| | sDHT type | | |
|---|---|---|---|
| | **Ambient**<br>**8 sDHTs** | **Wearable**<br>**29 sDHTs** | **Total**<br>**37 sDHTs** |
| **Data collection environment** | | | |
| *Actual environment; n* | 8 | 19 | 27 |
| *Simulated environment; n* | | 6 | 6 |
| *Both actual and simulated; n* | | 4 | 4 |
| **Interactions with sDHT** | | | |
| *Look-and-feel; n* | 1 | 3 | 4 |
| *Actual use; n* | 7 | 26 | 33 |
| **Human factors evaluation methods** [a] | | | |
| *Interviews; n* | 3 | 6 | 9 |
| *Focus groups; n* | | 2 | 2 |
| *Surveys - referenced; n* | 3 | 17 | 20 |
| *Surveys - in-house; n* | 4 | 13 | 17 |
| *Think-aloud; n* | 1 | | 1 |
| *Observation (direct or video); n* | | 11 | 11 |
| *Measured by the sDHT; n* | | | 0 |
| *Heuristic analysis; n* | | 2 | 2 |
| **Type/s of human factors data reported** | | | 0 |
| *Mixed methods; n* | 2 | 5 | 7 |
| *Quantitative only; n* | 4 | 20 | 24 |
| *Qualitative only; n* | 2 | 4 | 6 |
| **Categories of human factors and related data reported** | | | |
| *User satisfaction; n* | 5 | 23 | 28 |

| | | | |
|---|---|---|---|
| *Comfort; n* | 2 | 16 | 18 |
| *Ease of use; self-report; n* | 8 | 29 | 37 |
| *Ease of use; objectively-captured; n* | 1 | | 1 |
| *Learnability; n* | 1 | | 1 |
| *Efficiency; n* | | | 0 |
| *Memorability; n* | | | 0 |
| *Usefulness; n* | 5 | 19 | 24 |
| *Use errors; n* | 2 | 7 | 9 |
| *User trust; n* | 2 | 9 | 11 |
| *Readability; n* | | | 0 |
| *Understandability and/or actionability; n* | 1 | 5 | 6 |
| *Technical performance or product-errors; n* | 6 | 11 | 17 |
| **Adherence to sDHT reported** | | | |
| *Objectively-measured by the sDHT; n* | 2 | 7 | 9 |
| *Self/carepartner report; n* | 1 | 1 | 2 |
| *Both objective and self/carepartner; n* | | | 0 |
| *Reported but method not described; n* | | 1 | 1 |
| *Adherence not reported; n* | 5 | 20 | 25 |

[a] categories are not mutually exclusive
